# The Missing Link: A Closed Form Solution to the Kermack and Mckendrick Epidemic Model Equations

**DOI:** 10.1101/2021.03.02.21252781

**Authors:** Ted Duclos, Tom Reichert

**Author notes:** Drafts of this manuscript were edited by NeuroEdit.

## Abstract

Susceptible–infectious–recovered (SIR) models are widely used for estimating the dynamics of epidemics. Such models project that containment measures “flatten the curve”, i.e., reduce but delay the peak in daily infections, cause a longer epidemic, and increase the death toll. These projections have entered common understanding; individuals and governments often advocate lifting containment measures such as social distancing to shift the peak forward, limit societal and economic disruption, and reduce mortality. It was, then, an extraordinary surprise to discover that COVID-19 pandemic data exhibit phenomenology *opposite* to the projections of SIR models. With the knowledge that the commonly used SIR equations only approximate the original equations developed to describe epidemics, we identified a closed form solution to the original epidemic equations. Unlike the commonly used approximations, the closed form solution replicates the observed phenomenology and quantitates pandemic dynamics with simple analytical tools for policy makers. The complete solution is validated using independently measured mobility data and accurately predicts COVID-19 case numbers in multiple countries.

---

The susceptible–infectious–recovered (SIR) models (1), widely used in predictions of epidemic spread, are said to predict that social distancing will “flatten the curve”, i.e., reduce and delay the peak of new daily cases. These models project that the peak in daily cases will be reduced by a modest fraction, but the interval within which case numbers remain large lengthens in approximately inverse proportion. With this image highlighted in the popular media, concerns about the economic devastation projected to be associated with such measures applied in response to COVID-19 have caused many individuals and governments to advocate that social distancing measures to be lifted early (2) to shift the peak forward, to end the economic, educational, and social disruption sooner, and to reduce mortality.

As the COVID-19 pandemic swept across the world in the winter and spring of 2020, different countries applied diverse mitigation measures (3–5) in their attempts to control the spread of the virus. Although not deliberate, these countries have conducted “natural experiments” on the effectiveness of differing levels of containment measures. Their case data provides an historically unique opportunity to compare different models’ projections of the effect of diverse containment measures against real-world data.

In this manuscript, we first compare the trends in COVID-19 case data from countries that applied differing levels of containment to the trends predicted by an SIR model. We show that SIR models do not replicate even the basic trends in the country data. This discovery prompted us to develop a closed form solution to the original equations developed by Kermack and McKendrick (1) to which the SIR models are an approximation. We verify the solution by accurately predicting time series of case data for a sample of countries and by correlating the model to independent data. Finally, we show how to use the solution to manage epidemics with quantitative tools.

## EPIDEMIC MODELS

In 1927, Kermack and McKendrick (1) developed the following complex set of integro-differential equations for modelling epidemics:

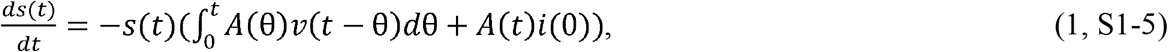

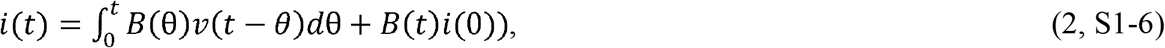

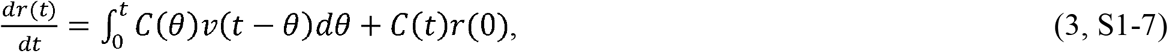

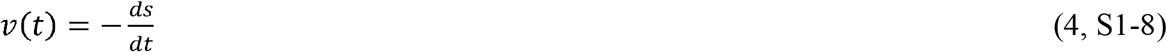

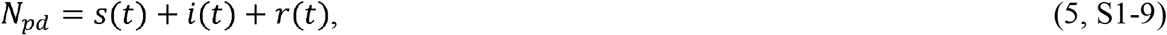

where 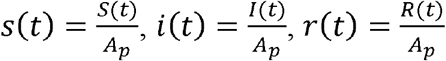, *S* = number of people susceptible to infection at time *t*; *I* = number of people infected at time *t*; *R* = number of people recovered at time *t*; *N*_*p*_ = total number of people in the population, 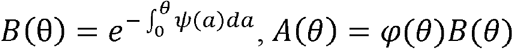, and *C* (*θ*) = *ψ* (*θ*)*B* (*θ*). In the original paper (1), *φ*(*t*) is defined as “the rate of infectivity”, *ψ*(*t*) is defined as “the rate of removal” of the infected population, and *N*_*pd*_ is defined as the initial population density. If *A*_*p*_ is defined as the area under consideration, then *N*_*pd*_*A*_*p*_ = *N*_*p*_, where *N*_*p*_ is the population under consideration. We call this set of equations the <u>C</u>omplete <u>SIR</u> (CSIR) model.

Although Kermack and McKendrick attempted to find an analytical solution for the CSIR equations, they, and many other researchers since (6), were only successful in finding solutions to approximations of their equations. One such set of approximate equations proposed by the original authors is shown here:

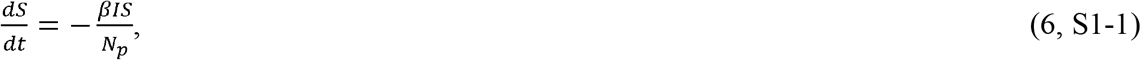

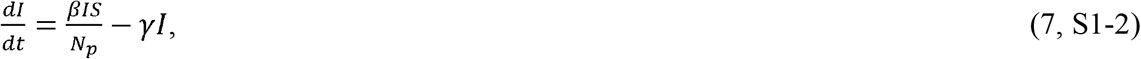

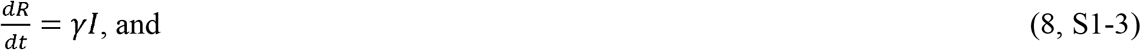

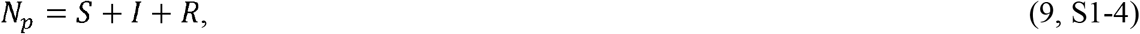

where *β* = rate of contact and transmission; *γ* = rate of recoveries = 1/*t*_*r*_; and *t*_*r*_ = time of infectiousness.

These equations (equations 6–9) are the well-known “SIR” model equations, which we will call here the <u>A</u>pproximate <u>SIR</u> (ASIR) model equations. These equations can be derived from equations 1–5 by assuming that the time-varying parameters, *φ*(*t*), and (*θ*) are the constants 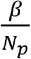 and *γ*, respectively. The ASIR model equations and their variants (SEIR, MSEIR, etc.) have been used for decades to model epidemics quantitatively and qualitatively.

A recently developed (7) analytical solution to equations 6-9 illustrates a problem with the ASIR equations. Within this solution, an expression depicting the time of the peak in infections increases as the value of *β* decreases until it becomes undefined in the region where 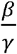 approaches 1. Although it is mathematically correct, this result casts doubt on the usefulness of the ASIR model since the condition, 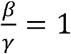, is the criterion for when the epidemic is beginning to end, and therefore, a useful model should be well-behaved at and in the vicinity of this condition.

## TESTING THE ASIR MODEL

Despite the mathematical problems with equations 6-9, the true test of a model, even an approximate one, is the extent to which it predicts real data trends. It is fortuitous, then, that the progression of the COVID-19 pandemic has been well documented in multiple countries which took different paths while attempting to contain the spread of the virus. This dataset affords us the opportunity to test the veracity of the trends predicted by ASIR model against actual data.

As a first step, we used equations 6-9 to project trends in the COVID-19 pandemic. In Figure 1A, we see that the ASIR model projects that the time at which the total number of cases levels off is delayed as social distancing increases (represented by decreasing *β*), while the ultimate total case number remains similar regardless of the value of *β*. In this simulation, increasing social distancing also delays and broadens the peak of new infections (Figure 1B).

**Figure 1.**
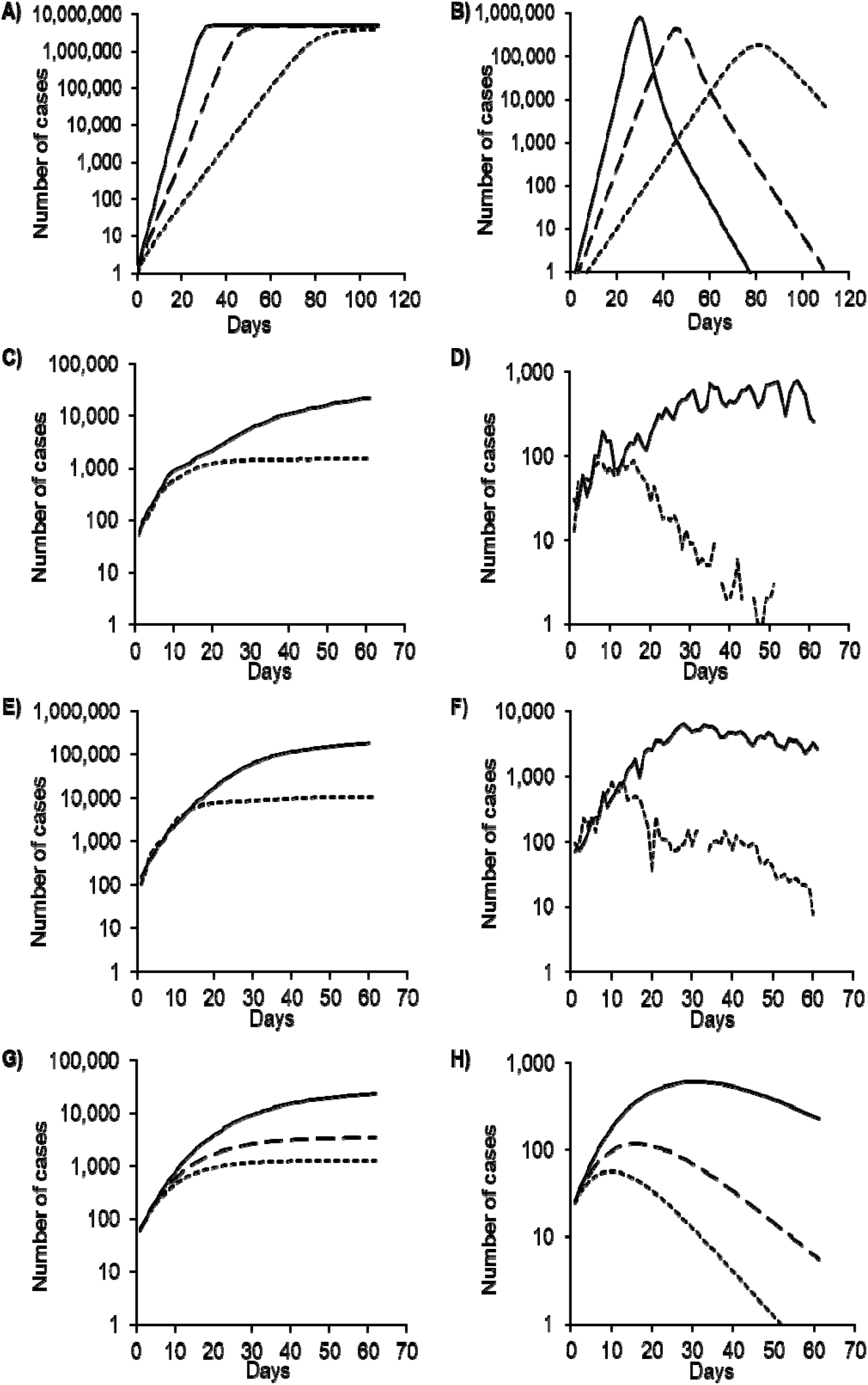
Comparisons of predictions of the approximate SIR (ASIR) and complete SIR (CSIR) models with observed data from four countries. Note: Containment measures increase in all panels from solid to long-dashed to short-dashed curves. **ASIR model trend predictions. (A)** Total cases; **(B)** Daily new cases. Rate of contact and transmission (β) decreases with increasing containment (from solid to dashed curves). Rate of recoveries (γ) = 0.2 for both plots. As β decreases, the daily total of cases increases more slowly and plateaus later (**A**). Daily new infections project to later, but only slightly lower peaks (**B**). Data reported from different countries during the COVID-19 pandemic Graphs are of data from pairs of countries with differing containment measures referenced to a day when each member of the pair had nearly equal numbers of new cases. **(C)** Total cases in Sweden (no containment measures, solid line) and New Zealand (strict containment, dashed line) **(D)** Daily new cases in Sweden (solid line) and New Zealand (dashed line) **(E)** Total cases in Italy (loose containment measures, solid line) and South Korea (strict containment, dashed line) **(F)** Daily new cases in Italy (solid line) and South Korea (dashed line) The trends in the observed data, panels (**C – F)**, are the opposite of those exhibited for increasing containment (decreasing β) in panels (**A, B**). CSIR model trend projections **(G)** Total cases; **(H)** Daily new cases. As containment measures increase (higher *K*_1_; solid to dashed curves), equation 18 projects that total cases will rise to lower levels, and reach these levels earlier **(G)** Similarly, equation 17 projects that new daily cases will peak earlier and lower values with increasing containment **(H)**. The CSIR model trends in **(G)** and **(H)** are highly similar to those in the country data **(C – F)**. The ASIR model trends in (**A**) and (**B**) have completely different shapes; and vary with increasing containment in an opposite sense to that in the country data.

In a second step, we compared the ASIR projected trends to case data (8) for the COVID-19 pandemic in Sweden and New Zealand (Figure 1C, D), and for South Korea and Italy (Figure 1E, F). These pairs of countries have comparable population densities, but implemented mitigation measures with different timings and intensities (3–5, 9). In particular, New Zealand and South Korea introduced stronger social distancing measures much earlier than Italy and Sweden.

In contrast to the ASIR model predictions, the country data in Figures 1C–F show that stronger social distancing measures are associated with an <u>earlier</u> and <u>lower</u> peak in new infections and an <u>earlier</u> levelling off at a <u>lower</u> total number of cases. Both trends demonstrate that the ASIR model (Figure 1A, B) is not merely inaccurate but projects epidemic data to trend in the *opposite* direction to the reported data. Other authors (10), too, have noted that the peaks of cases in countries with stronger social distancing occur earlier than in countries with weaker social distancing.

The ASIR model fails the simplest test of model veracity: the projection of qualitative trends. The model simply does not match real-world experience and all variants of the ASIR model suffer from this fault.

## A SOLUTION TO THE CSIR MODEL EQUATIONS

Since ASIR equations fail to capture the true dynamics of real-world experience, we revisited the original CSIR equations developed by Kermack and McKendrick to determine if either an analytical solution or new approximation that preserves the underlying dynamics could be found. In Supplement 1, we derive an exact closed form solution to the original Kermack and McKendrick model equations. This solution was developed by first recasting the original equations into a new set of differential equations, and then solving these equations using basic principles. The closed form solution incorporates measurable population interactions, and can be manipulated using straightforward mathematical techniques to reveal a number of relationships useful for managing an epidemic.

As a first step towards a solution, the CSIR equations (equations 1–5) were recast into the following set of differential equations:

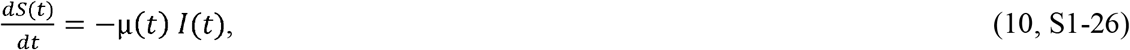

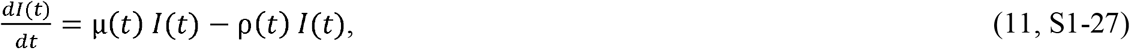

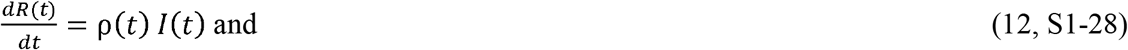

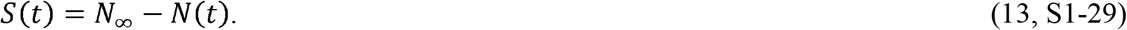

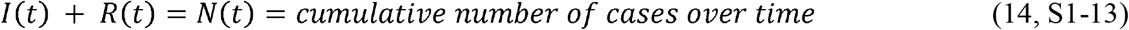

using these definitions for µ (*t*)and ρ(*t*):

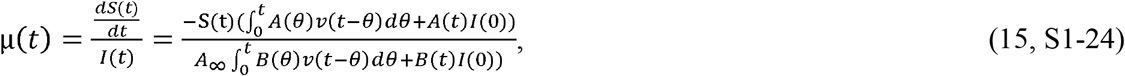

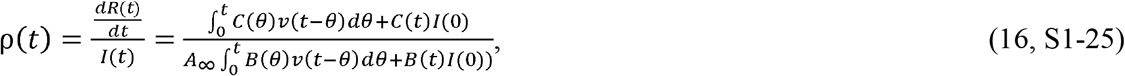

where 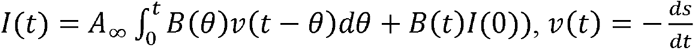, and *A*_∞_ is the area that the population *N*_∞_ inhabits.

As detailed in Supplement 1, equations 10–16 were developed by adopting a perspective from inside the epidemic rather than using an external view. Equations 10-16 are also shown to be mathematically identical to equations 1–5 in Supplement 1, but recast in a form for which a solution can be found.

In a second step, using basic principles (Supplement 1) and equations 10, 13 and 14, we developed the following differential equation to describe the new daily cases during an epidemic:

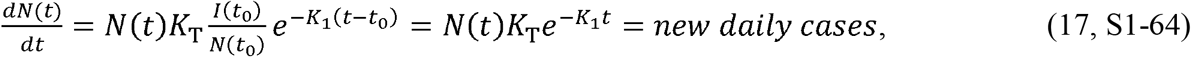

where *K*_T_ is a constant representing the disease transmissibility, 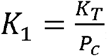, and *P*_*c*_ is the number of specific people a person contacts in an infectious manner during the time period 0–*t*.

The solution to equation 17 is:

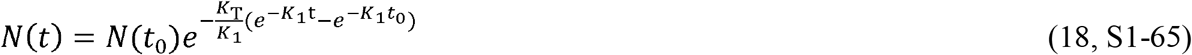

and, in Supplement 1, the following are shown to be the solutions to equations 10, 11 and 12:

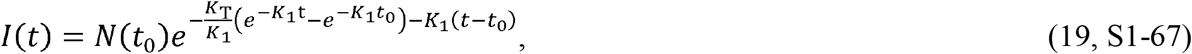

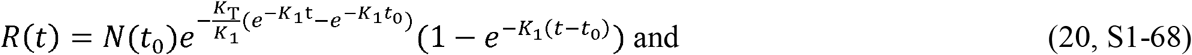

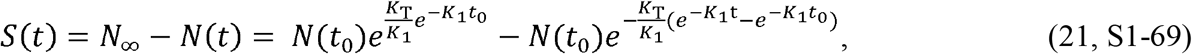

where:

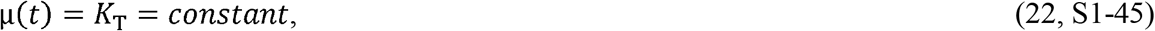

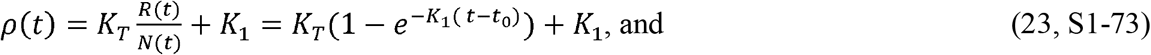

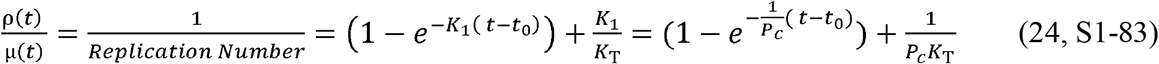

Equations 18–24 are a closed form solution to the original model proposed by Kermack and McKendrick and, as we will show in the next section, they accurately describe the evolution of an epidemic.

## VERACITY OF THE CSIR SOLUTION

The qualitative behavior of the CSIR solution is plotted in Figure 1G, H. The CSIR solution predicts that with increased social distancing (higher *K*_*1*_), the total number of daily cases is lower and attained earlier (Figure 1G). The peak in new cases is also lower and occurs earlier (Figure 1H). Unlike the ASIR model trends (Figure 1A, B), the CSIR solution trends (Figure 1G, H) are identical to those exhibited by the real-life COVID-19 data in Figure 1C–F. Whereas the ASIR model fails to predict the proper impact of social distancing, the CSIR solution predicts the trends seen in the real data and, in particular, that stronger social distancing produces an earlier peak of daily cases. We also note here that, relative to the native, unmanaged epidemic, for all containment measures (higher *K*_*1*_), the peak is lower and occurs earlier. In other words, the curve never flattens; rather, it reaches a peak earlier and then falls in a steep decline.

Qualitative projection is a low bar for model validation. A useful model must also make accurate predictions. To demonstrate the predictive capability of the CSIR solution, we tested it quantitatively on data from several countries by first estimating two country-specific constants, *K*_1_ and *K*_2_ (where *K*_2_ = ln (*K*_*T*_)) These constants were found by calculating a measure called the rate of change operator (RCO) (equation S1-81). This operator linearizes equation 17 and enables the determination of the parameters *K*_1_ and *K*_2_ for each country.

Since the resulting RCO time series (Figure 2) all have distinctive segments where the curves appear to become straight lines shortly after intervention measures were imposed in each country, the parameters *K*_1_ and *K*_2_ can be determined by finding the slopes and intercepts of these lines. Values for *K*_1_ and *K*_2_, tabulated in Table 1, were estimated by fitting a linear expression to a small early portion of these straight segments. These early portions were nine data points in length and their date ranges are shown in Table 1. The derived values for *K*_1_ and *K*_2_ were then used to predict the balance of the epidemic data that followed.

**Figure 2.**
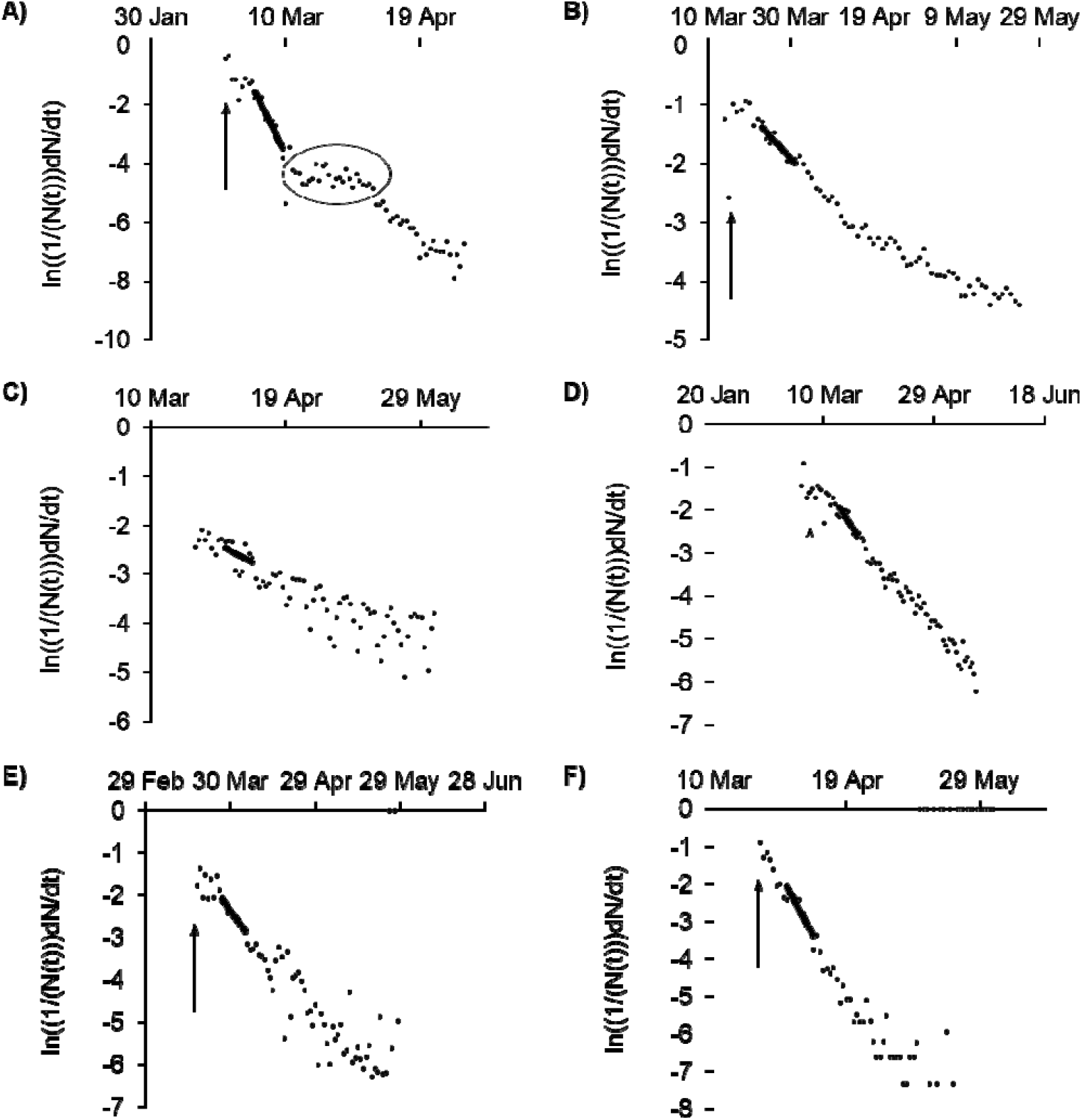
Rate of change operator (RCO) curves for tCOVID-19 cases in various countries. **A)** South Korea (the oval highlights a departure of the observed data from the slope, indicating failures in, or relaxations of, social distancing); **B)** USA; **C)** Sweden; **D)** Italy; **E)** Spain; and **F)** New Zealand. An epidemic can be described by a piecewise linear model using the RCO (equation S1-6). A short solid line on each graph shows a line segment fit to the corresponding points in the observed data. The slopes and initial points of these line segments are the values of and respectively which are tabulated in Table1. RCO curves change markedly soon after the date containment measures were implemented (arrows: South Korea, February 21; USA, March 16; Italy, March 8; Spain, March 14; New Zealand, March 25. Sweden did not implement any specific containment measures, so the model calibration was begun on April 1, when the slope of the rate of change operator (RCO)curve first became steady.). All dates are in 2020.

**Table 1.**
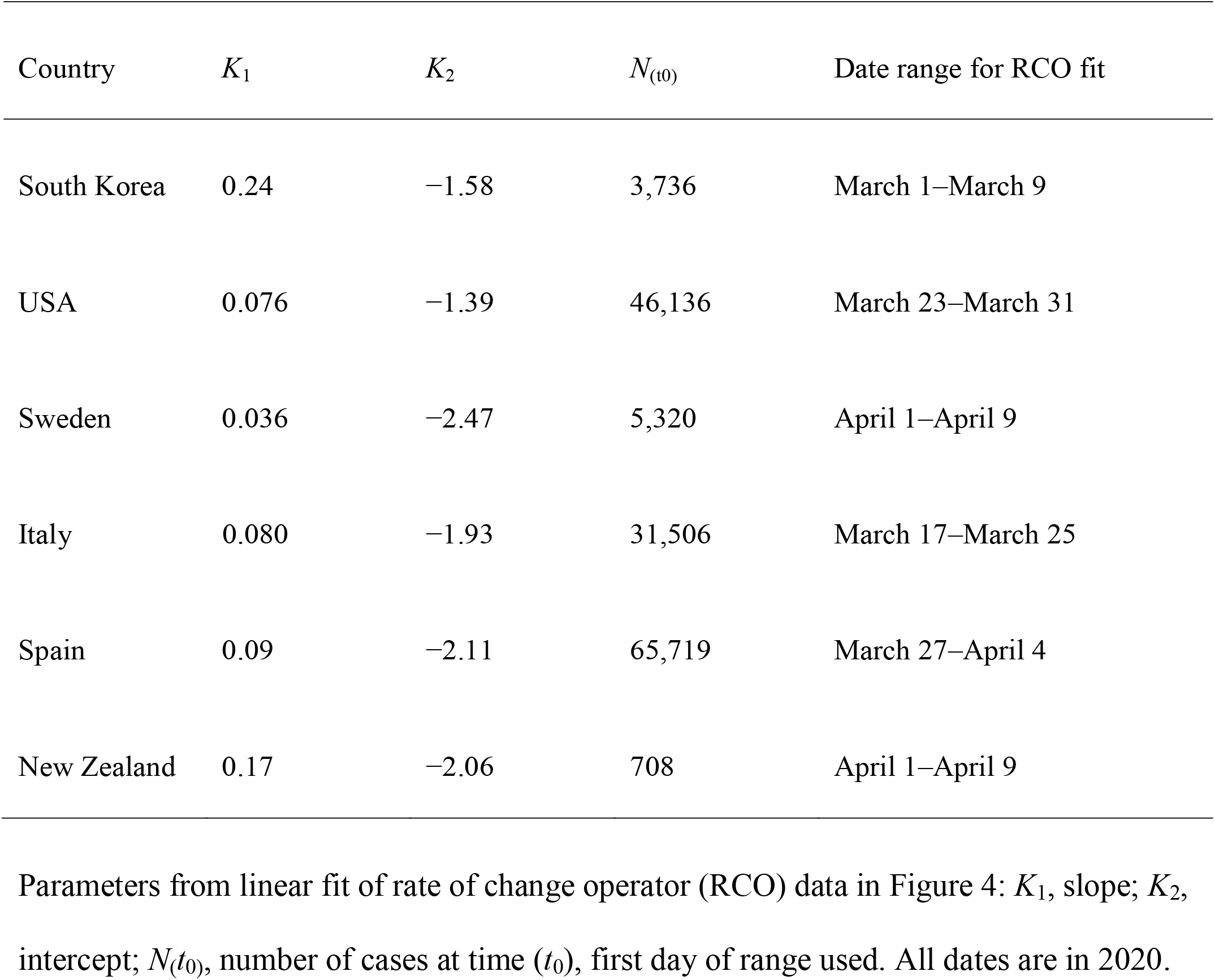
Social containment parameters used to model total cases and new daily cases of infection.

| Country | $K_1$ | $K_2$ | $N_{(t_0)}$ | Date range for RCO fit |
| --- | --- | --- | --- | --- |
| South Korea | 0.24 | −1.58 | 3,736 | March 1–March 9 |
| USA | 0.076 | −1.39 | 46,136 | March 23–March 31 |
| Sweden | 0.036 | −2.47 | 5,320 | April 1–April 9 |
| Italy | 0.080 | −1.93 | 31,506 | March 17–March 25 |
| Spain | 0.09 | −2.11 | 65,719 | March 27–April 4 |
| New Zealand | 0.17 | −2.06 | 708 | April 1–April 9 |

Using this approach, the CSIR solution predicted the course of total cases (Figure 3) with an *R*^*2*^ > 0.97 for all six countries for the 45 days following the date containment measures were introduced (Table 1). In the case of Sweden, which did not introduce containment measures, March 23, 2020 was chosen as the starting point.

**Figure 3.**
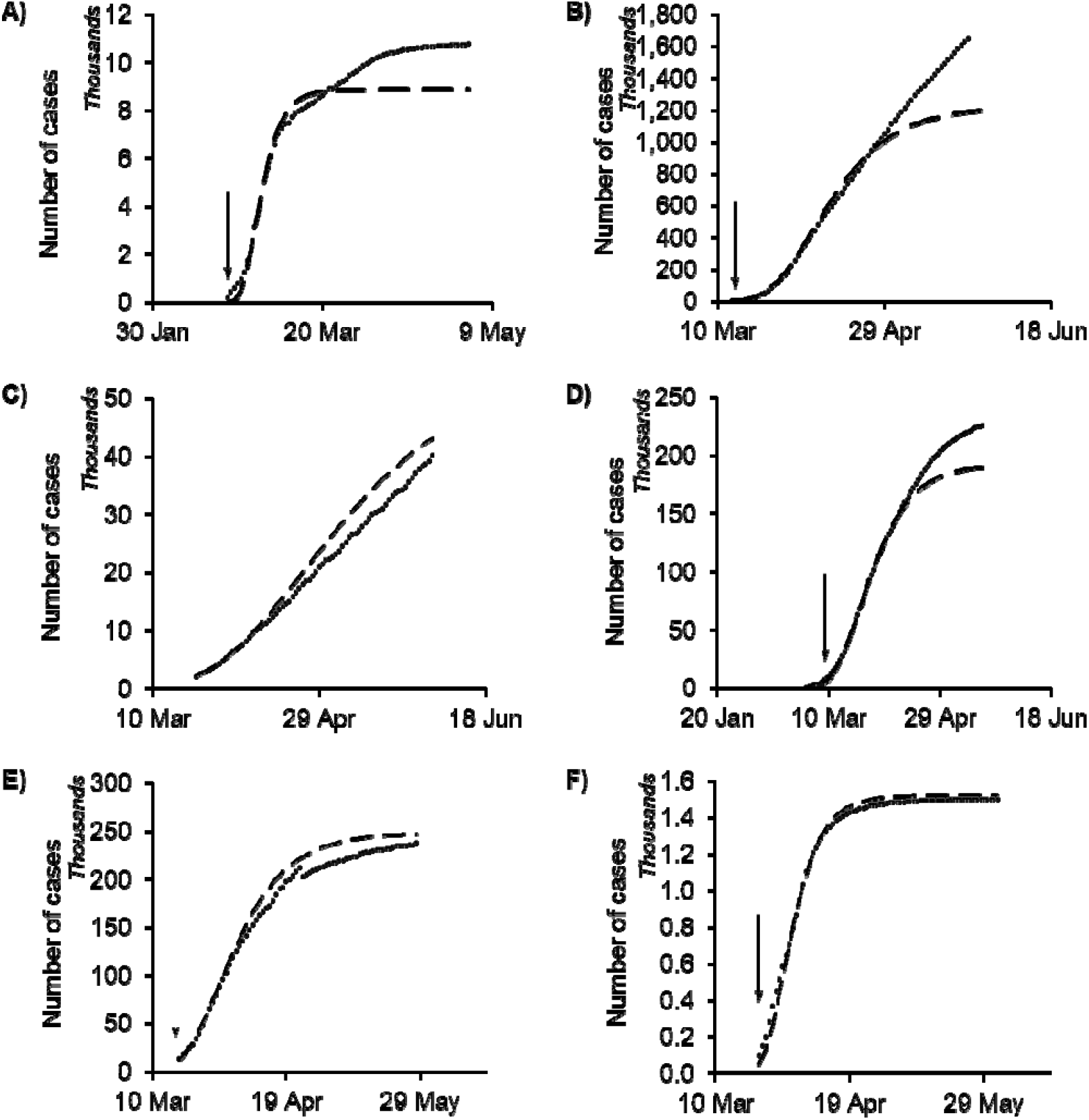
Complete SIR (CSIR) model predictions for daily total case counts. **A)** South Korea; **B)** USA; **C)** Sweden; **D)** Italy; **E)** Spain; and **F)** New Zealand. Dashed line, model; dots are individual data points, observed from each country. *R*^2^ > 0.97 for all countries for 45 days after the containment measures were implemented: South Korea, February 21-April 4; USA, March 16-April 30; Italy, March 8–April 22; Spain, March 14-April 28; New Zealand, March 25-May 9. Sweden did not implement any specific containment measures, so the dates used were March 23-May7. The CSIR model was calibrated using data from the date ranges listed in Table 1. The poor correlation of the model to the data in the USA panel (**B**) after April is explained in Supplement 2. All dates are in 2020.

The CSIR solution also predicted daily new cases (Figure 4) for these six countries for the 45 days following the date containment measures were introduced with an *R*^*2*^ range of 0.29 to 0.90 As seen in the figure, the predicted peak of new cases was close to the observed peak for all countries.

**Figure 4.**
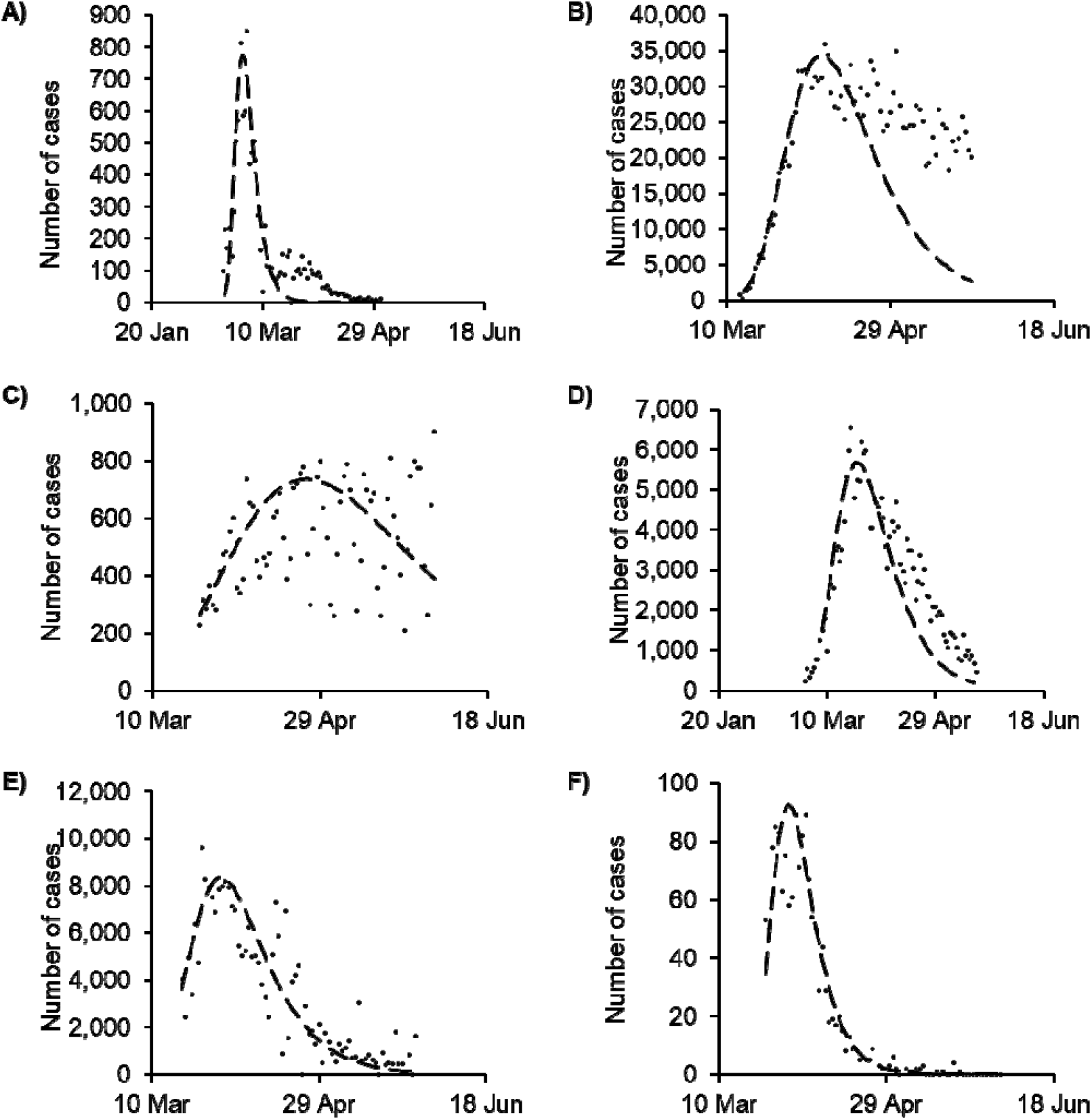
Complete SIR (CSIR) model predictions for number of new daily cases. **A)** South Korea, *R*^2^ = 0.86; **B)** USA, *R*^2^ = 0.83; **C)** Sweden, *R*^2^ = 0.29; **D)** Italy, *R*^2^ = 0.69; **E)** Spain, *R*^2^ = 0.65; and **F)** New Zealand, *R*^2^ = 0.90. Dashed line is the model in all panels; dots are data points, observed daily from each country. *R*^2^ values are between the model and the data across countries for the 45 days after containment measures were imposed. All dates are in 2020.

It is important to emphasize that the CSIR projections in Figures 3 and 4 are *not* fits to the full-length of the data shown. Rather, a short portion of the epidemic data, with specified characteristics, was excised for the purposes of determining coefficients of the model; these were then used to project the rest of the curves before and after the time frame of the excised data.

In an additional demonstration of the CSIR solution’s veracity, we tested the assumption that *K*_*T*_ is a property of the disease; and, therefore, should be the same for each country. Equation S1-79 shows that the model parameters, expressed in a function, *F(N(t))*, should be linearly proportional to time with a constant of proportionality −*K*_*T*_. As illustrated in Figure 5, the fit of equation S1-79 to the country data from Table 2 has an *R*^*2*^ = 0.97 and a slope of −0.25 (which is equal to −*K*_*T*_). This excellent correlation confirms that *K*_*T*_ is the same for all countries; and, therefore, likely is a property of the disease.

**Figure 5.**
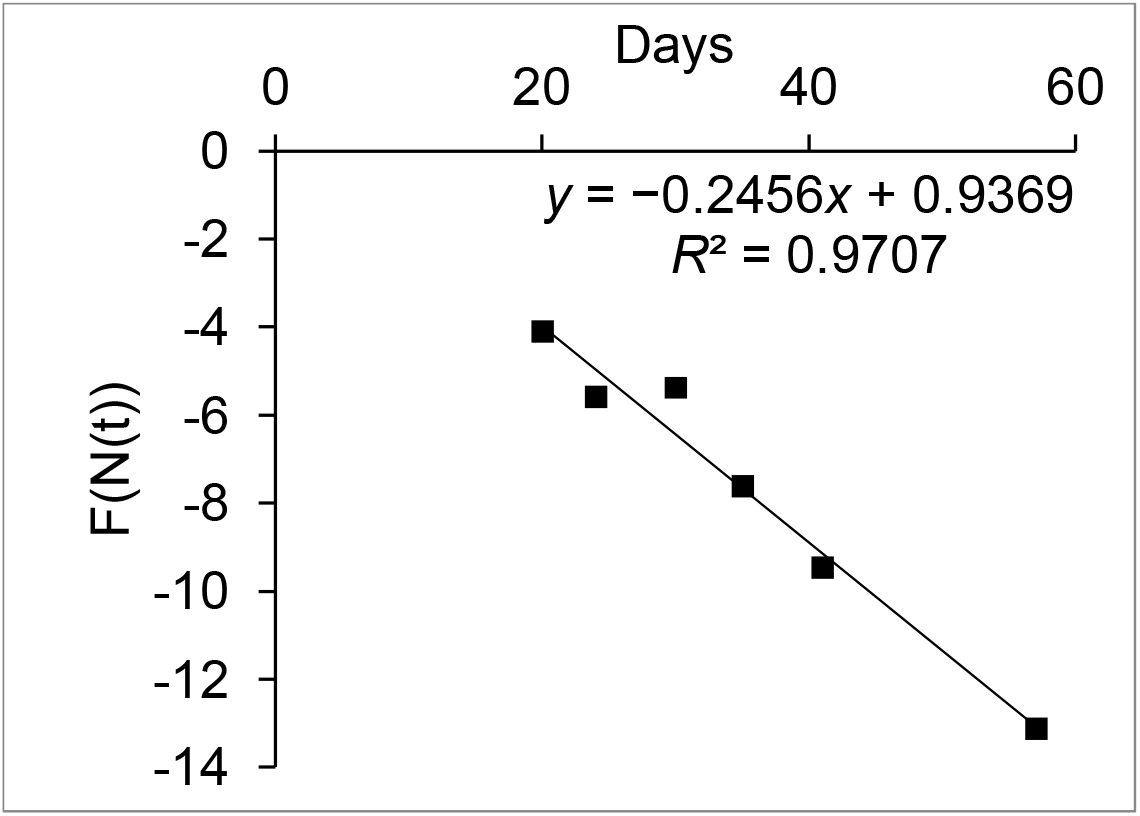
Verification that *K*_*T*_ is the same for all countries. The data from Table 2 is plotted using equation S1-79 and *A*_1_ = 0.53 *km*^2^. Each data point is a value corresponding to a different country. The value of *K*_*T*_ is the negative of the slope of the line, and *K*_*T*_ is closely approximated everywhere by *K*_*T*_ ≈ 0.246.

**Table 2.**
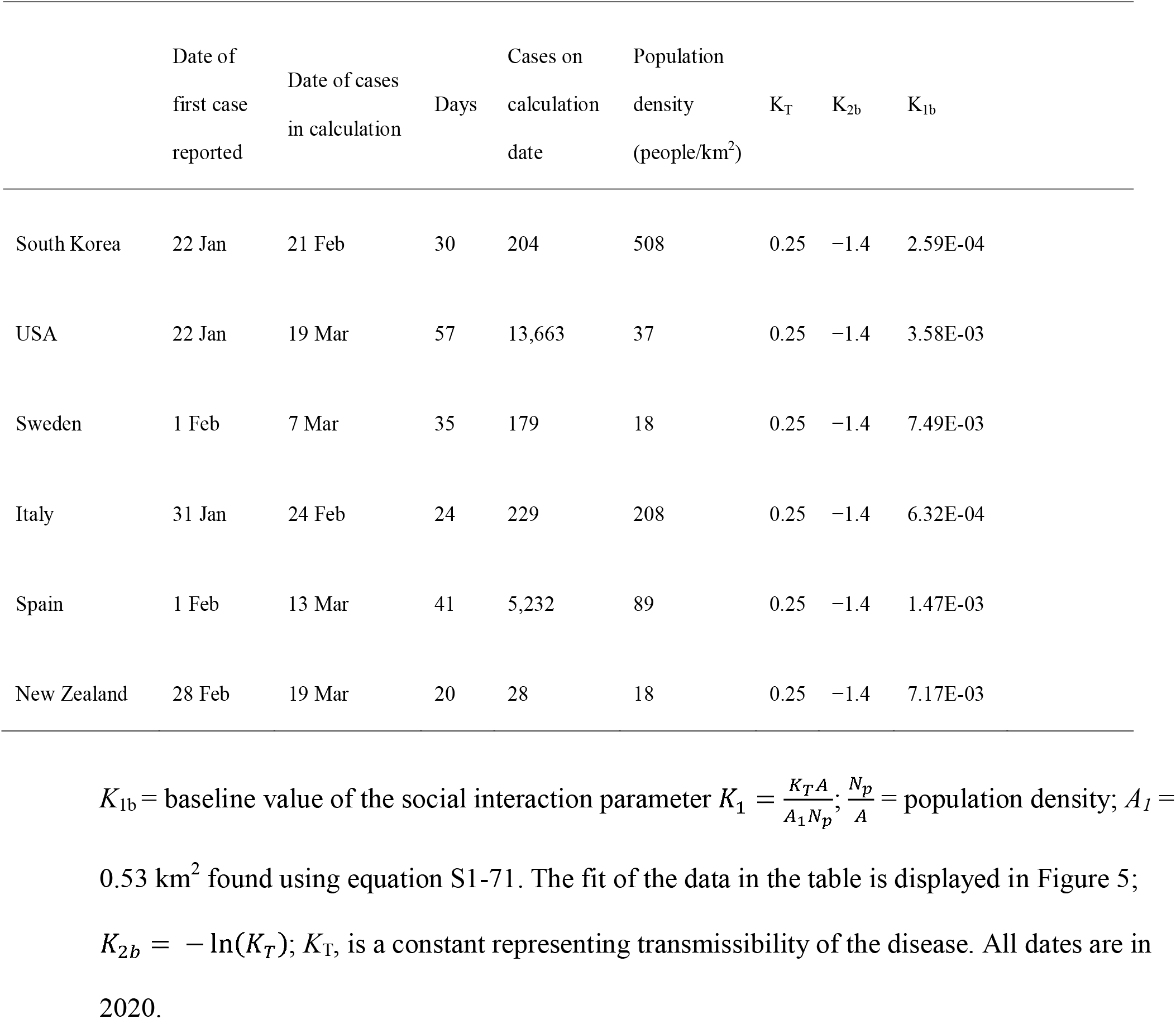
Initial COVID-19 pandemic data and social interaction parameters for various countries

| | Date of<br>first case<br>reported | Date of cases<br>in calculation | Days | Cases on<br>calculation<br>date | Population<br>density<br>(people/km<sup>2</sup>) | $K_T$ | $K_{2b}$ | $K_{1b}$ |
| --- | --- | --- | --- | --- | --- | --- | --- | --- |
| South Korea | 22 Jan | 21 Feb | 30 | 204 | 508 | 0.25 | -1.4 | 2.59E-04 |
| USA | 22 Jan | 19 Mar | 57 | 13,663 | 37 | 0.25 | -1.4 | 3.58E-03 |
| Sweden | 1 Feb | 7 Mar | 35 | 179 | 18 | 0.25 | -1.4 | 7.49E-03 |
| Italy | 31 Jan | 24 Feb | 24 | 229 | 208 | 0.25 | -1.4 | 6.32E-04 |
| Spain | 1 Feb | 13 Mar | 41 | 5,232 | 89 | 0.25 | -1.4 | 1.47E-03 |
| New Zealand | 28 Feb | 19 Mar | 20 | 28 | 18 | 0.25 | -1.4 | 7.17E-03 |

$K_{1b}$ = baseline value of the social interaction parameter $K_1 = \frac{K_T A}{A_1 N_p}$ ; $\frac{N_p}{A}$ = population density; $A_l =$
0.53 km<sup>2</sup> found using equation S1-71. The fit of the data in the table is displayed in Figure 5;
$K_{2b} = -\ln(K_T)$ ; $K_T$ , is a constant representing transmissibility of the disease. All dates are in 2020.

A third illustration of the solution’s veracity was created by correlating mobility data obtained over the modelled period for each of the countries. These data are available from Google (11) and are a measure of the difference between the amount of time people stayed at home (the “Residential” data set) during the period modelled and a baseline measured for 5 weeks starting January 3, 2020. Based on equation S1-46, the integral of this mobility data should correlate with the measured RCO. Figure 6 shows that for each country considered, the correlation was quite high.

**Figure 6.**
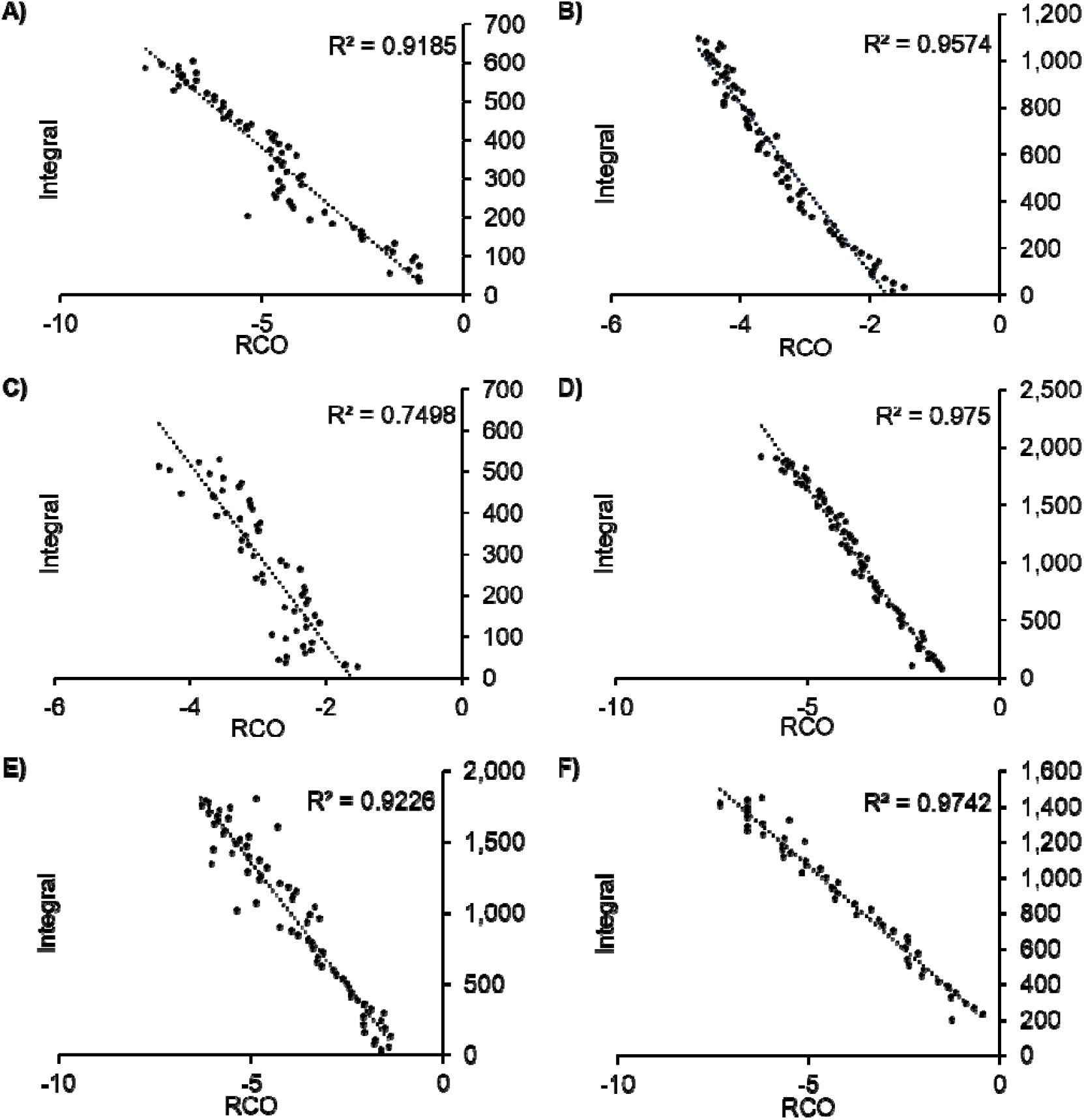
Correlations between the daily value of the rate of change operator (RCO) and the integral of the Google Residential Mobility data. **A)** South Korea (date range, February 23 to April 23); **B)** USA (March 25 to May 31); **C)** Sweden (March 5 to May 5); **D)** Italy (March 25 to May 31); **E)** Spain (March 25 to May 31); and **F)** New Zealand (March 21 to April 22). All dates are in 2020.

These excellent results demonstrate that the CSIR solution, embodied by equations 18–24, correctly characterizes epidemic dynamics from multiple countries in a unified way, something that ASIR models simply cannot duplicate. They also demonstrate that the model developed by Kermack and McKendrick nearly 100 years ago, described by equations 1-5 and recast as equations 10-16, was correct and accurately captures the dynamics of epidemics.

## USING THE SOLUTION TO CONTROL EPIDEMICS

Many important relationships and identities can be derived from equations 18–24. They are too numerous to be listed here; therefore, they are described and explained in detail in Supplements 1, 2, and 3. Specifically, in Supplements 1 and 2 we develop identities that illustrate the importance of early intervention strategies. Subsequently, in Supplement 3, we derive mathematical tools that can be used by public health officials to characterize, control, and end an epidemic.

As explained in Supplement 2, the importance of early and strong intervention cannot be overstated. The growth of the infected population, *N*(*t*) (equation 18) is doubly exponentially dependent on the behavior of the population. This means that early and strong social distancing actions will end the epidemic early, reducing the number of cases in an exponential fashion. The peak in daily cases will never be delayed by strong actions but will, in fact, come earlier. The curve does not flatten with strong intervention. It steepens and falls to an early end.

If an epidemic is not controlled in its early stages, the tools explicated in Supplement 3 can be used to determine the state of the epidemic and to calculate quantitative actions that must be taken to control and end the epidemic. Strong intervention actions are still necessary and, using the RCO metric, the effectiveness of these actions can be quickly determined from the resultant case trends. The epidemic state, tracked with the RCO metric, can be closely monitored and control measures adjusted according to the observed trends.

As an epidemic progresses, outbreaks and surges should be expected. These will occur if containment flags, more infectious variants emerge, or new cases are introduced from outside the region of focus. All will need to be controlled. As explained in Supplement 3, the start of an outbreak causes obvious changes in the behavior of the RCO metric. Diligently monitored, these RCO changes can be detected early enough for public health officials to react in a timely manner, to bring outbreaks under control, and to re-establish the planned course.

## DISCUSSION

The data on the current COVID-19 pandemic are widely accessible from a variety of sources, updated daily. The unfolding panorama provides a test bed for models used to predict outcomes and the effects of various interventions. Because different countries have employed different containment strategies (3-5, 9), the world is conducting an epidemiological experiment on a grand scale.

Current epidemiological models use the ASIR equations, which have been assumed to be reasonably accurate representations of the complete equations developed in 1927. They exist in many variants, both deterministic and stochastic; and their behavior is widely known. It is startling, then, that when the classic ASIR model is tested using the currently available data from the COVID-19 pandemic, it fails the most basic test for any model: it projects trends that are the opposite of those easily visible in the data from many countries.

Unfortunately, the ASIR model and its variants have been used to fashion guidelines for epidemic management. Tragically, the false notion that stronger social distancing lengthens the pandemic may well have caused country leaders, especially those most concerned with economic performance, to see social distancing as producing only a modest reduction in the horror of an epidemic peak while significantly prolonging economic disruption.

The CSIR solution, developed from basic mathematical principles, accurately projects the epidemic trends. It makes clear that short and sharp social distancing produces rapid truncation of epidemic upward trends, thereby shortening—not lengthening—the time needed to bring epidemics under control. Second, an indicator of the rate of change in epidemic dynamics (the RCO) allows direct observation of the effectiveness of intervention measures and provides policy makers with an opportunity to react before new outbreaks gain momentum.

Every country and economy can use the solution presented here to plan and implement the highest level of social distancing measures deemed sustainable to quickly reduce case numbers to levels at which case identification, contact tracing, testing, and isolation can be maintained, allowing a more rapid return to nearly normal social interactions while minimizing economic consequences. The ultimate insight from the model is one of hope: the path of an epidemic is not an uncontrollable force of nature; nor is epidemic control inevitably the road to economic ruin. Rather, the afflicted population can, through their behavior, choose to control their destiny.

## Data Availability

All data on the country cases is available at: 8. Roser M, Ritchie H, Ortiz-Ospina E, et al. Coronavirus pandemic (COVID-19). Our World in Data. https://ourworldindata.org/coronavirus. Published February 2, 2021. Accessed February 2, 2021.
Data on mobility is available at: 11. Google mobility reports. https://www.google.com/covid19/mobility/. Published December 24, 2020. Accessed December 24, 2020

https://ourworldindata.org

https://www.google.com/covid19/mobility/

## SUPPLEMENTARY MATERIALS

### SUPPLEMENT 1

[Equations are numbered sequentially. Main text numberings appear in bold.]

The <u>A</u>pproximate <u>SIR</u> (ASIR) model

The SIR model, with non-time-varying parameters *β* and *γ*, is described by the following equations:

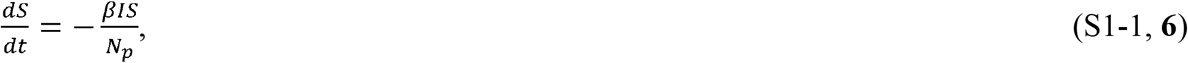

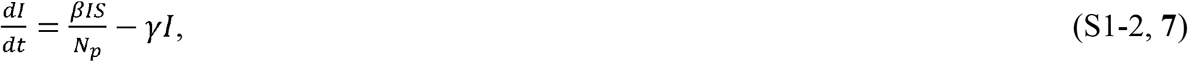

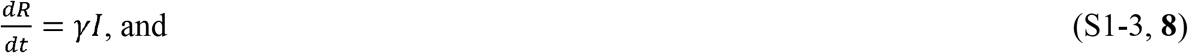

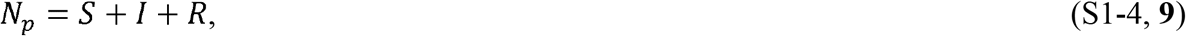

where *S* = number of people susceptible to infection at time *t*; *I* = number of people infected at time *t*; *R* = number of people recovered at time *t*; *N*_*p*_ = total number of people in the population; *β* = rate of contact and transmission; *t*_*r*_ = time of infectiousness; *γ* = rate of recoveries = 1/*t*_*r*_. This formulation is from Kermack and McKendrick (1) and is widely used to demonstrate the qualitative behavior of the more complex model with time-varying parameters presented in the original paper.

A solution to equations 1–4 is easily obtained using straightforward computational methods. Figure 1A and 1B depict one such solution computed using a simple Euler method to solve these differential equations with a time step of 1 day.

Because the assumption that *β* and *γ* are constant strongly limits the capability to describe epidemics, we call this form of the model the “approximate SIR” (ASIR) model. While not intended to model epidemic dynamics perfectly, equations 1–4 have always been assumed to reflect the general trends of an epidemic (1). As Figure 1 clearly demonstrates, however, these equations do not even qualitatively reproduce the dynamics of an epidemic.

#### The <u>C</u>omplete <u>SIR</u> (CSIR) model

Referring to the original equations (1), we can write the epidemic equations without any assumptions about the time-varying nature of any parameter, using our notation of S, I, and R from above as:

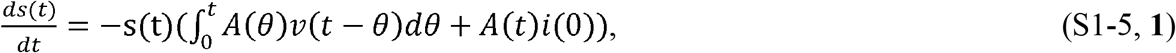

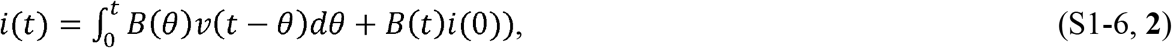

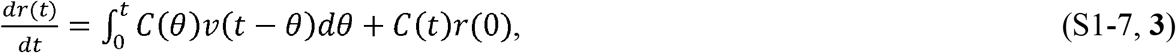

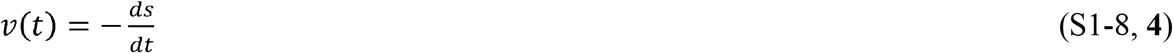

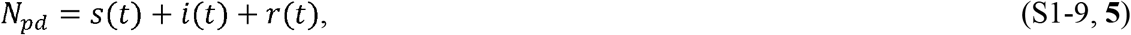

where 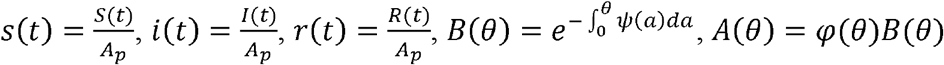 and *C(θ) = ψ*(*θ*) *B*(*θ*).*φ*(*t*) is defined as the rate of infectivity at age *θ* ”, *ψ*(*t*) is defined as “the rate of removal” of the infected population, *N*_*pd*_ is defined as the initial population density, and *A*_*p*_ is defined as the area under consideration. Therefore, *N*_*pd*_ *A*_*p*_ = *N*_*p*_, where *N*_*p*_ is the population under consideration. Equations 1–4 can be derived from equations 5–9 if the quantities *φ*(*t*) and *ψ*(*t*) are assumed to be the constants 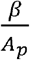 and *γ* respectively, and equations 5–9 are all multiplied by *A*_*p*_.

For the development of our solution, we apply a new perspective to equations 5–9. Rather than look at the epidemic as involving the total population, *N*_*p*_, we will focus on only that portion of the population that will eventually become infected in the epidemic. We call this subpopulation *N*_∞_. We also introduce a further subpopulation of *N*_∞_ which we call *N*_*S*_*(t)* and which we define as the population at time *t* that is under consideration and is in contact with the epidemic. This mchange in perspective views the epidemic mathematically from inside the bounds of the already infected population which is ever expanding rather than mathematically describing what is happening within a fixed total population.

The reinterpretation begins by first recognizing that, in equations 5–7, the initial number of infections introduced to the population—which we will designate *I*_*i*_—is merely the starting value of the epidemic and can be any value at all. The second step is to imagine that at every time, *t* the epidemic starts again and the initial infections introduced into the population, *I*_*i*_(*t*), are equal to the current infections. The third step is to recognize that the values of the integrals in equations S1-5–7 are equal to zero and *B*(*θ*) whenever the epidemic starts. With these perspectives in mind, if equations S1-5–7 and S1-9 are first all multiplied by *A*_*S*_ the area that *N*_*S*_(*t*) inhabits when *t* = 0), they can be rewritten as

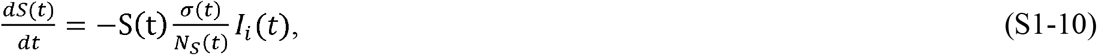

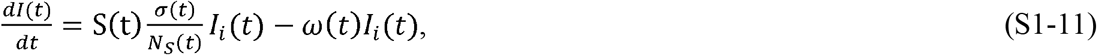

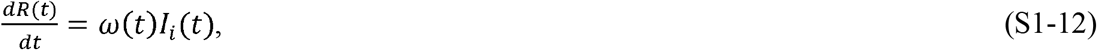

and since there are only susceptible people in *N*_*S*_(*t*) when the new infections are introduced,

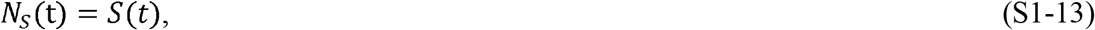

where *σ*(*t*) = *φ*(*t*)*N*_*S*_(*t)* and *ω*(*t*) = *ψ*(*t*).

If we also now define a quantity, *R*_*i*_(*t*), as the number of recovered persons introduced to the population at the same time as *I*_*i*_(*t*), we can write an equation for *N*_∞_ and define a new quantity, *N*(*t*), the number of people either currently infected or recovered,

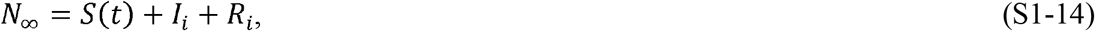

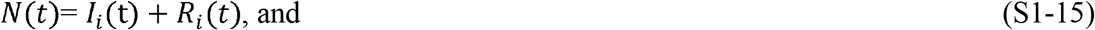

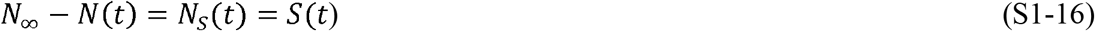

We have not assumed that any of the quantities are constant; therefore, we can now explicate the time-varying nature of these quantities. We begin by considering what happens to equations S1-10–12 during the time interval Δ*t* from a time *t* to *t* +Δ*t*, and by rewriting these equations as difference equations:

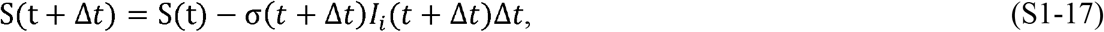

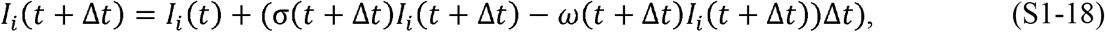

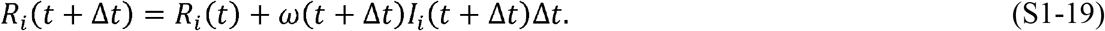

Taking the limit as Δ*t*→0, we obtain the following differential equations:

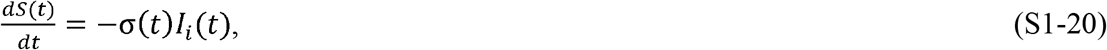

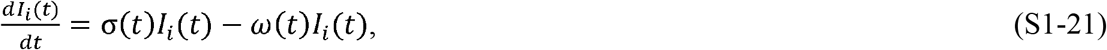

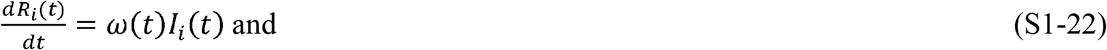

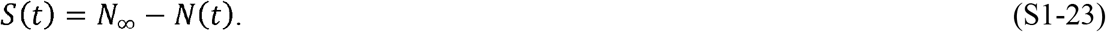

From the preceding, it can be easily seen that *I*_*i*_(*t*) = *I*(*t*) and *R*_*i*_(*t*) = *R*(*t*), so for the remainder of the analysis, the subscript, *i*, will be dropped.

The perspective in the immediately preceding part of the analysis is that the epidemic can be considered to start over again at each instant in time. In this way, the susceptible population is not fixed by initial conditions, but rather is the population that will eventually become infected in the future. Embedded in this concept is the assumption that, during each Δ*t*, the susceptible population is always in contact with those people that have been previously infected or who will become infected, i.e., the epidemic remains contiguous. This is not restrictive when considering the initial stage of the epidemic; however, it plays a critical role in recognizing an outbreak that starts outside the population, *N*_∞_.

Equations S1-20–23 can also be developed from equations S1-5–7 using a different consideration. If we first define two functions μ(*t*)and ρ(*t*)as

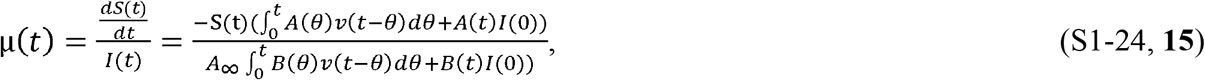

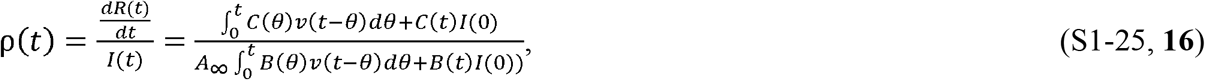

then we can rewrite equations S1-5–7 in the following form after multiplying by*A*_∞_:

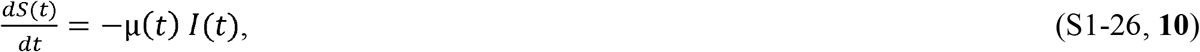

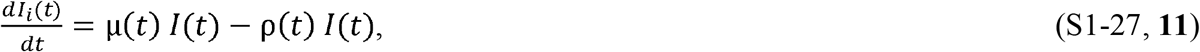

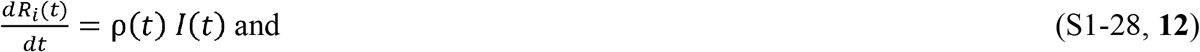

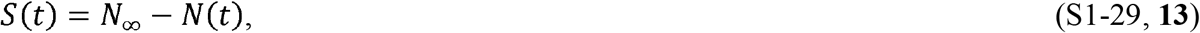

where 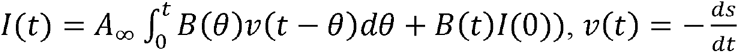, and *A*_∞_ is the area that the population *N*_∞_ inhabits.

Comparing equations S1-26–29, we can see that if we set μ(*t*) = σ(*t*) and ρ(*t*) = ω(*t*), then equations S1-26–29 are also equivalent to equations S1-20–23. Both sets of equations are mathematically equivalent to the Kermack and McKendrick (1) system of equations, S1-5–9.

We have recast the equations by shifting the population and area under consideration to *N*_∞_ and *A*_∞_, respectively, rather than *N*_*p*_ and *A*_*p*_, and defined the susceptible portion of the population during the epidemic as those that will eventually become infected under the conditions in place during each instance in time. However, this shift in perspective retains the mathematical equivalence to the equations derived by Kermack and McKendrick (1) because the portion of *N*_*p*_ that is not a part of *N*_∞_ never becomes infected and therefore never affects the values of *I*(*t*)nor *R*(*t)* in the solutions to equations S1-5–9 or S1-26–29.

#### The solution to the CSIR model

We now need to find a solution to equations S1-26–29. We begin by reiterating the definitions of *I*(*t*), *R*(*t*), and *N*(*t*): *I*(*t*) are individuals who are currently infectious, *R*(*t*) are individuals who have had the infection but are no longer contagious, and *N*(*t*) is the sum of these individuals, i.e., *N*(*t*) = *R*(*t*) + *I*(*t*). The population, *R*(*t*), is also referred to as the recovered population because although they may still be ill, they are no longer infectious.

We assume that people in the recovered population remain immune after recovery and that the epidemic begins with the introduction of the infection by one individual at time *t=* 0; therefore, *I(0) = N(0) = 1*. In addition, we assume that no new infections (other than the initial infection) are introduced from outside the region of interest during the epidemic.

The goal in solving equations S1-26–29 is to find an expression for *N*(*t*) and for the total number of people who will be infected during the epidemic, defined as *N*_∞_, in terms of the parameters of disease transmission and the behavior of the population enduring the epidemic. We will first find an expression for *N*(*t*) and then find *N*_∞_ by taking the limit of *N*(*t*) as time increases. Along the way, we will also find expressions for *I*(*t*), *R*(*t*), and *S*(*t*).

The development of the solution begins by noting the following:

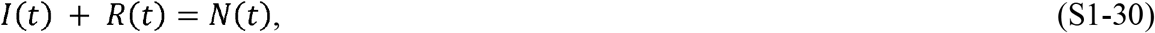

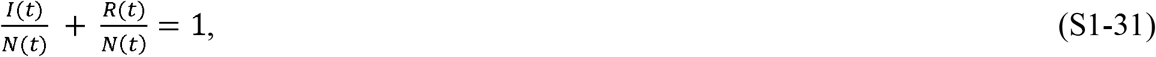

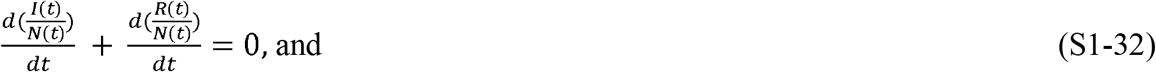

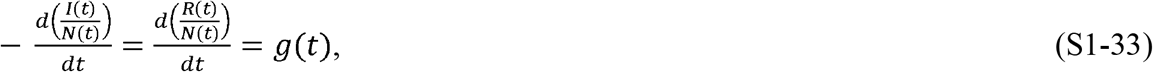

where *g*(*t*) is a yet unknown function of time.

We need an expression for the function *g* (*t*) in terms of the variables and parameters defined thus far. We know that 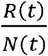 can only change if *I* (*t*) changes, since all the recovered people must have been infected; therefore, 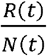 can only change to exactly the extent that the quantity 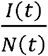 changes. Thus, we can write the following expression:

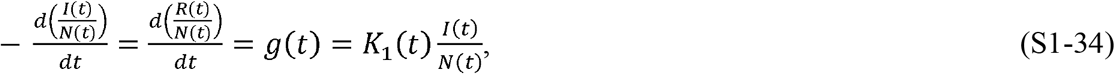

Where *K*_1_(*t*) is a function that modifies the fraction 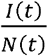 in association with recoveries at time *t*. It is initially assumed to be a function of time.

Equation S1-34 is a simple differential equation in the variable 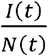, whose solution is

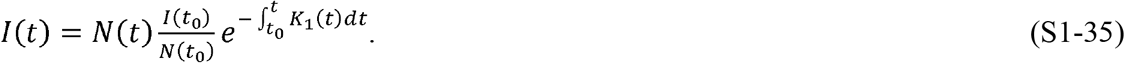

Or, if *K*_1_(*t*) is a constant,

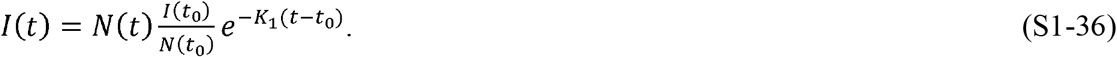

Likewise, the solutions for *R*(*t*) are

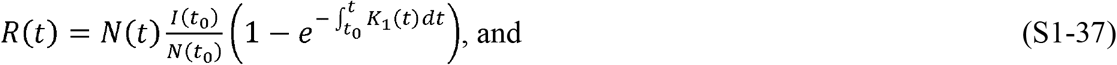

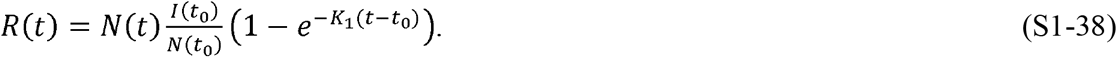

When time *t*_0_ = 0, *N* (*t*_0_)= *I*(*t*_0_) =1 and equations S1-35–38 become

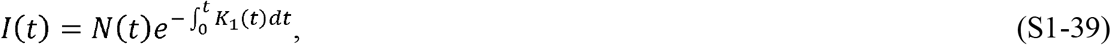

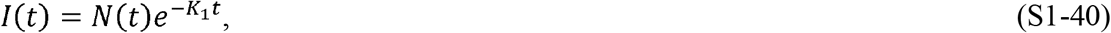

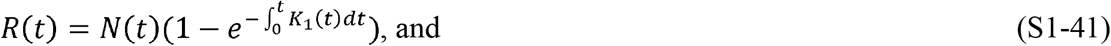

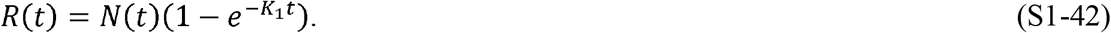

During an epidemic, if every contact made by a person within the population *N*(*t*) could result in an infection, then the disease progression can be represented by the following expression:

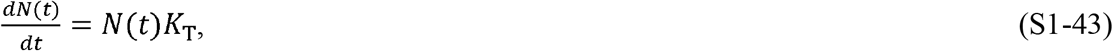

where *K*_T_ is a constant representing the transmissibility of the disease.

Because *N*(*t*) consists of both infected and recovered individuals, we can rewrite equation S1-43 as

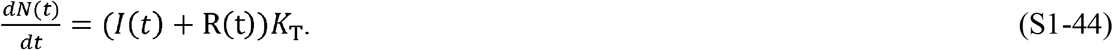

Because the population *R*(*t*) cannot transmit the disease, *R*(*t*) *K*_T_ = 0, equation S1-44 can be rewritten as

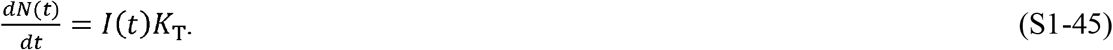

Since 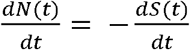, equation S1-45 implies that *K*_T_=μ(*t*) and that μ(*t*) is, in fact, a constant.

Furthermore, this means that 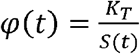.

Using equation S1-45 and equations S1-35 or S1-36, we can write

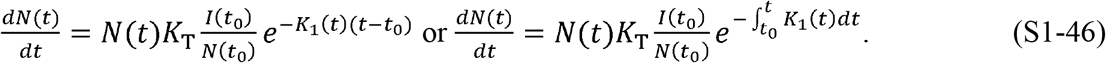

Both forms of equation S1-46 can be solved for *N*(*t*), but before we do this, we will elucidate the physical meaning of *K*_1_ to improve our understanding of the solution.

We begin by defining a new parameter,*Pc* (*t*):

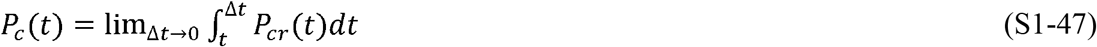

where *P*_*cr*_ (*t*) is contact rate for the subpopulation *N* (*t*).

*P*_*c*_ (*t*) can be interpreted as the average number of specific infectious contacts each person within the subpopulation *N*(*t*) has within the entire population. “Specific infectious” is intended to clarify that each person is assumed to interact only with the same people (quantity = *P*_*c*_(*t*)) in a way they might possibly transmit the disease during the time under consideration. Based on this definition, the number of interactions with infectious potential between the population *N*(*t*) and the entire population is *N*(*t*)*P*_*c*_(*t*), at time *t*.

For simplicity in the expressions going forward, we define *F*_*i*_(*t*) as the fraction of *N*(*t*) that is infected:

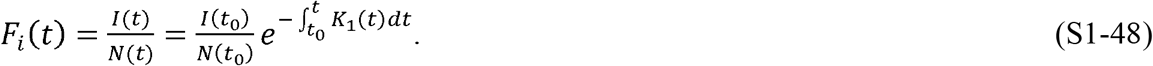

Using the definitions immediately above, we can write the following expression using equation S1-46:

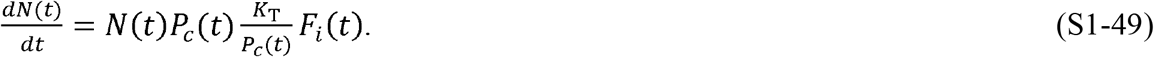

Equation S1-49 can then be written as the following difference equation:

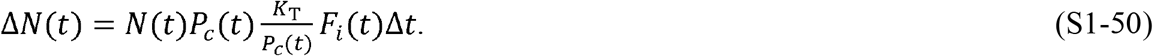

Equation S1-50 can also be rewritten in the following manner:

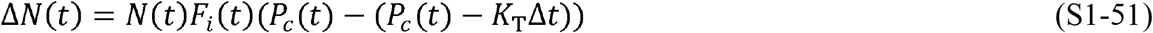

or

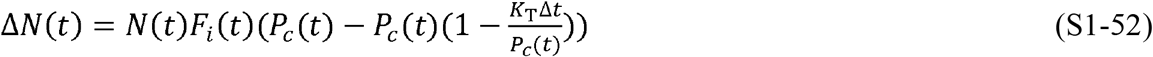

In equation S1-51, the term (*P*_*c*_(*t*) − *K*_T_Δ*t*) is the change in infectable contacts in the time Δ*t* due to infections that occurred during time Δ*t*. Here, we define “infectable contacts” as the number of contacts within *P*_*c*_ (*t*) that might become infected, because they have not been infected previously. Likewise, the term 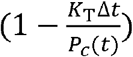 in equation S1-52 is the change in the fraction of infectable contacts during Δ*t*. Since *P*_*c*_(*t*) is independent of the infectable contacts, the change in the fraction of infectable contacts must be the fraction that *F*_*i*_ (*t*) changes during Δ*t*. Rewriting equation S1-52, we can express this mathematically as

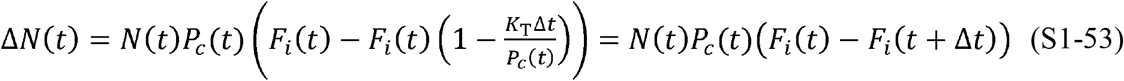

Equation S1-53 can be used to develop a recurrence relationship to explain the nature of *K*_1_.

At *t* = 0, *F*_*i*_ (0) = 1and equation S1-53 becomes

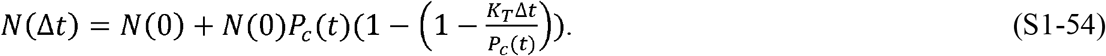

We should also note that

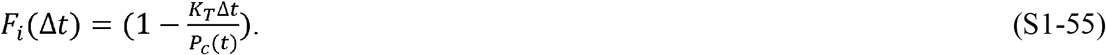

In the next time step, applying equation S1-52 to equation S1-54, we find the following expression:

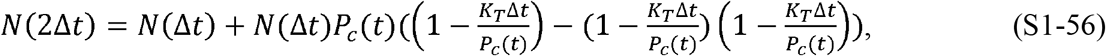

noting that 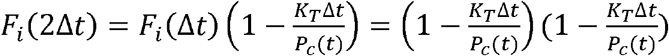.

Equation S1-56 can be simplified to

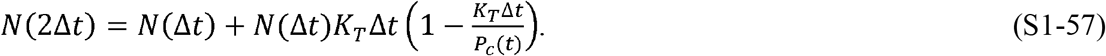

Repetitive application of the same logic leads to the recurrence relationship

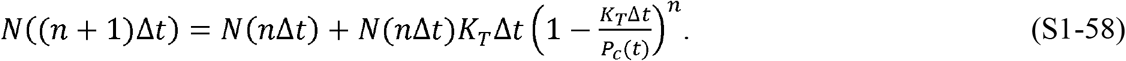

Rearranging terms and defining *n*Δ*t = t*, we can write the following:

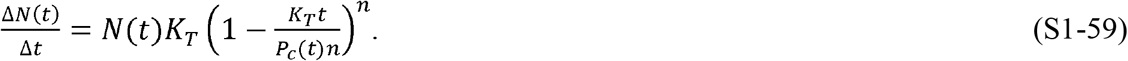

If we now allow Δ*t*→0, and therefore, *n* → ∞, and note that 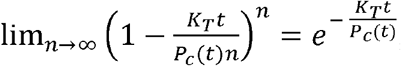, we obtain the following expression for 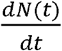:

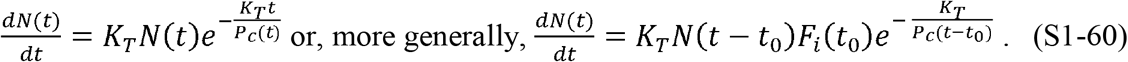

Comparing equation S1-60 to equation S1-46, we can see that 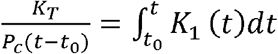; or, if *P*_*c*_(*t*) is a constant,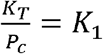 and 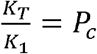.

Returning to *P*_*c*_(*t*), we will now further define this parameter. As previously defined, *P*_*c*_(*t*) is the number of specific infectious contacts a member of the population *N*(t) has within the entire population and is a function of the population’s behavior. Initially, we assume this to be a function of population density. We define this area as the effective area rate,*A*_1*r*_(*t*). Using these assumptions, we can write an expression for *P*_*cr*_(*t*):

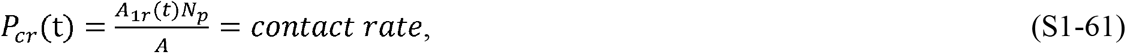

where *N*_*p*_ = the population of the region with the infection, *A*= the area of the region, and 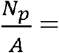 the population density. From equation S1-61, we can see that *P*_*cr*_(*t*) is proportional to both the population’s behavior, *A*_1*r*_(*t*), and the population density, 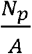.

Similar to the way we defined *P*_*c*_(*t*) using *P*_*cr*_(*t*), we can define *A*_1_(*t*) in terms of *A*_1*r*_(*t*):

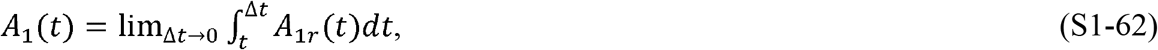

Where *A*_1_(*t*)is the effective specific area traversed by a person. In this case, “specific” has the same meaning as it has for *P*(*t*)_*c*_ : each person traverses only and exactly the same area for the duration of the time under consideration. We also call *A*_1_(*t*)the “effective area” because the population is typically only dispersed within ∼1% of the land in a given region within a country (12). If we take this into account, then 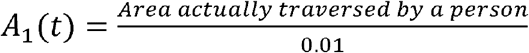.

From the preceding discussion, we can now write an expression for *K*_1_(*t*):

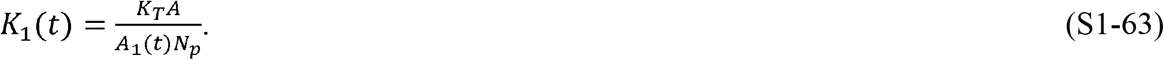

Equation S1-63 shows that, under the initial assumptions of immunity, no cases from outside, and a contiguous epidemic, *K*_1_ > 0. Since *K*_1_(*t*) is inversely proportional to *A*_1_(*t*), as people reduce the area they traverse per unit time by increasing social distancing or taking other measures, *K*_1_(*t*) increases due to the lowered contact rate. Therefore, *K*_1_(*t*) is inversely proportional to the strength of social distancing interventions implemented during an epidemic.

The physical meanings of *P*_*c*_(*t*) and *A*_1_(*t*) may not be immediately intuitive. As defined, neither are rates, but they can both vary in time and their values are dependent on the population’s behavior. They are, respectively, the number of specific infectious people contacted and the specific area traversed by an index person within a given time. They are constant when the number of specific infectious people or the traversed area remain constant. However, if different people are contacted within a given time period, the rates they are dependent upon will change, and therefore, *P*_*c*_(*t*) and *A*_1_(*t*) will change even if the total number of people contacted or area covered has not changed during that time period.

Since both the population’s behavior and the population density represented by *A*_1*r*_(*t*) and 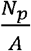 are likely to remain constant for many days running, for the initial model development, we can safely assume that *P*_*cr*_(*t*) is constant and, therefore, that *P*_*c*_(*t*) and *K*_1_(*t*) are constants. Of course, we only expect this to be true on a piecewise basis because, eventually, in response to the epidemic, populations take measures such as mask wearing, online meetings, quarantines, and re-openings, which significantly change the contact rate and therefore *P*_*c*_(*t*). The piecewise variation of *K*_1_(*t*) is addressed later in a separate section.

Having defined *K*_1_(*t*)as a piecewise constant, for simplicity, we will refer to it as *K*_1_and to *P*_*c*_(*t*) as *P*_*c*_ for the remainder of this portion of the analysis. We can now find the solution to equations S1-26–28 and S1-5–7.

Using the expression for 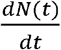 from equation S1-46, we arrive at the following:

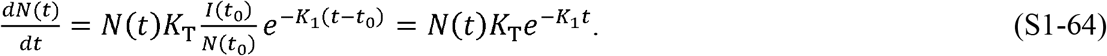

Integrating S1-64 produces the expression for the total number of infections, *N*(*t*) :

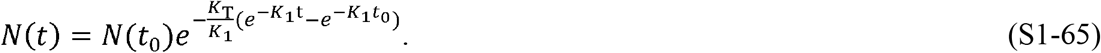

Taking the limit of equation S1-65 as, *t* →∞, we obtain the expression for the total number of individuals who will be infected within the entire epidemic:

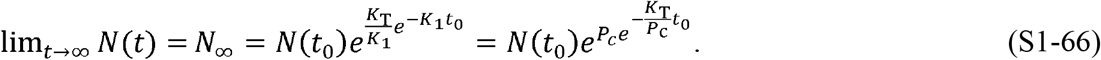

We can write expressions for *I*(*t*) and *R*(*t*) as

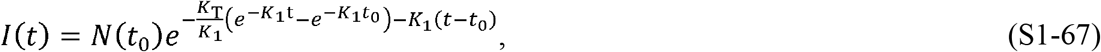

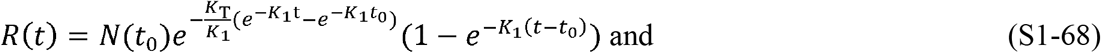

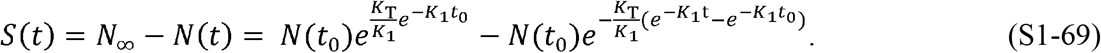

Equations S1-67–69 can each be differentiated and rearranged to produce these differential equations:

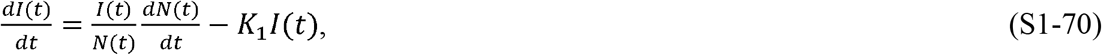

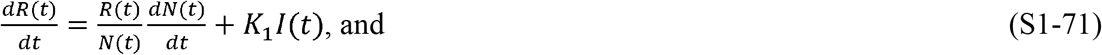

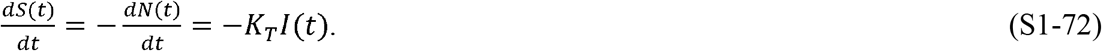

As previously discussed, *K*_T_ = μ(*t*). If we also equate equations S1-71 and S1-12, we can find an expression for *ρ*(*t*) :

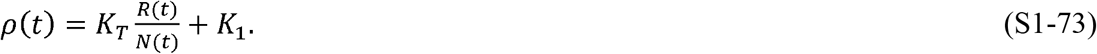

Using these definitions for *μ*(*t*) and *ρ*(*t*), we can see that equations S1-70–72 are equivalent to equations S1-26–28 and equations S1-5–7. Therefore, equations S1-67–69 are solutions to the time-varying equations in the CSIR model proposed nearly 100 years ago by Kermack and McKendrick.

#### Insights into the CSIR solution

Equations S1-66 and S1-69 introduce an important and rather abstract concept, not previously discussed. Recall that our starting point was a reset of perspective to allow the size of the susceptible population to be variable. The susceptible population is the subpopulation of the total population that *will become* infected. Equation S1-69 describes precisely how the susceptible population changes as the behavior of the population, 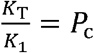, changes. The equation shows that the susceptible population size is not a pre-ordained constant and it quantifies the manner in which the susceptible population can rise or fall, independent of the change in infections.

Mathematically, equation S1-66 demonstrates that the number of infections to be expected, *N*_∞_, and the behavior of the population, *P*_*c*_, are interrelated in the sense that changes in *P*_*c*_ have a doubly exponential effect on the final number of infections that will come to be. This underscores that small changes in population behavior dramatically affect the epidemic’s outcome. However, the reciprocity means that the eventual number of cases produced by the epidemic is also not foreordained but rather a strong function of interventions introduced.

We gain additional insight into the meaning of the CSIR model solution by looking at equation S1-70 in more detail. Equation S1-45 can be used to rewrite equation S1-70 as

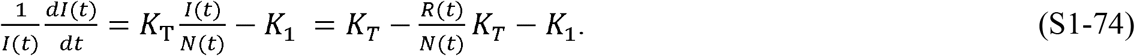

The left-hand side of equation S1-74 is the rate of change in the number of new infections per person currently infected. The first two terms on the furthest right-hand side of equation S1-74, 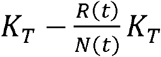, describe the rate of successful infection. These terms account for contacts by the recovered population in a way that ASIR models do not. Furthermore, since *K*_*T*_ is the rate at which an infected person causes infections per infectable contact, the terms 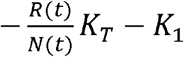 must represent the rate of recovery per infected person.

One more insight into the solution is gained from the following relationship, derived from equations S1-64–66:

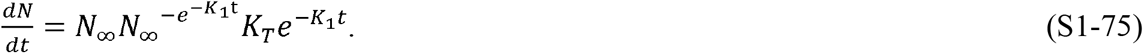

In words, the form of equation S1-75 is

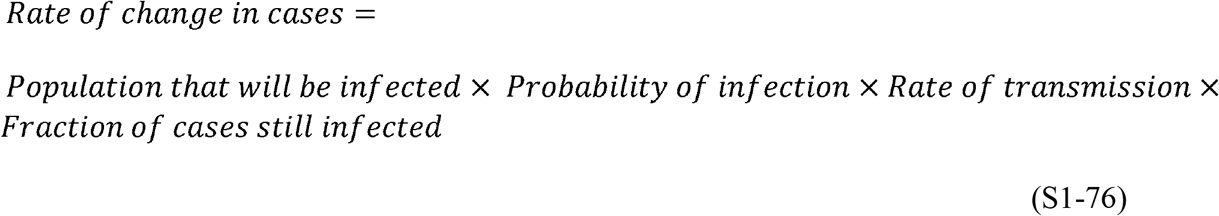

or

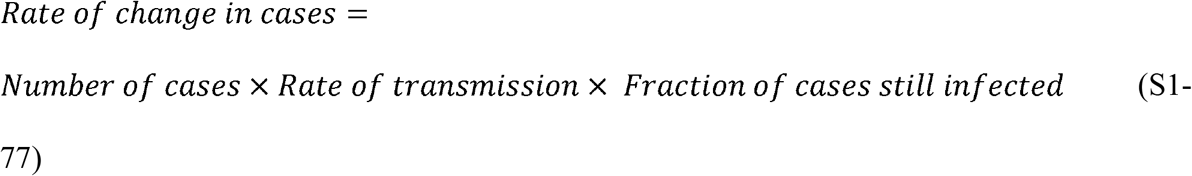

Equations S1-76 and S1-77 illustrate the logic of the CSIR solution in terms of probabilities.

Finally, using the prior definition, *F*_*i*_(*t*) =*Fraction of cases still infected*, and if *t*_0_ = 0, we can use equation 65 to write this simple expression for the CSIR solution:

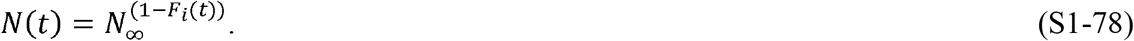

#### Model verification

The solution to equations S1-5–9 was developed assuming that *K*_*T*_ is a constant dependent only on the disease, and *K*_1_ that is dependent on the population behavior. These assumptions were used in generating the correlations of the case data for various countries in Figures 2 and 3. However, both assumptions can be tested independently from the country correlations.

The check of the assumption that *K*_*T*_ is a constant can be made by substituting equation S1-63 into equation S1-65 and solving for *K*_*T*_*t*. Doing this, we find the following expression:

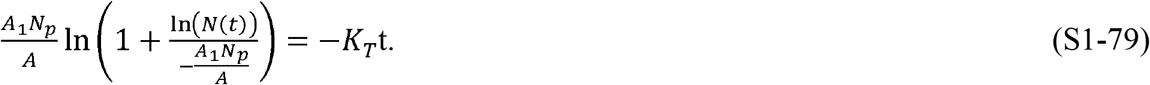

If we define 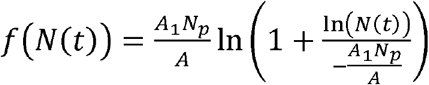, then we can also write this expression:

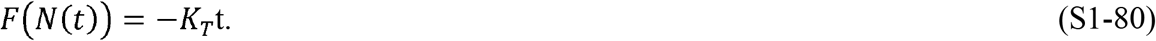

If *K*_*T*_ is a constant, then equation S1-80 predicts that *F* (*N*(*t*)) is a linear function of time. Excepting *A*_1_, all the quantities on the left-hand side of equation S1-70 can be derived from the country data. Therefore, we can estimate the value of *K*_*T*_ using equation S1-79 (or S1-80) to find the value of *A*_1_ that best fits a straight line. This fit is illustrated in Figure 5 and shows a correlation coefficient of 0.955, which strongly supports the assumption that *K*_*T*_ is a constant.

Equation S1-64 can be rewritten to define an important relationship:

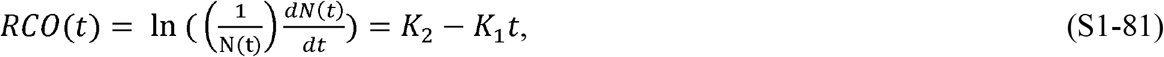

where 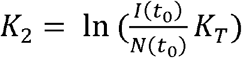 and *t*_0_ is the time between the start of the epidemic and the time at which the first data point in the epidemic is measured.

The RCO expression is convenient in that it transforms equation S1-64 into an equation linear in *K*_1_. We call this expression the “rate of change operator” since it is the rate of change of new cases scaled by the current daily case number. Equation S1-81 is the equation that was fit to the country data in Figure 2, and used to find the parameters in Table 1. Note that if *K*_1_ decreases and if it approaches zero, the growth of the epidemic will be nearly exponential.

Similarly, we can use the fact that equation S1-46 predicts that the RCO measure is proportional to 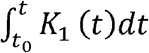, where *K*_1_ is a measure of the interactive behavior of the population. If an independent measure of people’s mobility during the epidemic should also be linearly related to the RCO, we can have additional confidence in the veracity of the CSIR solution. Google has compiled different measures of people’s mobility derived from mobile phone data (11). One of these measures is termed the residential mobility measure (RMM). The RMM is a measure of the percent increase or decrease that people stayed in their residence during the pandemic relative to a baseline measured over 5 weeks starting on January 3, 2020. Since *K*_1_ and the RMM are both inversely proportional to the population’s mobility, the RMM should be a good proxy for the value of *K*_1_. To test this, we plotted the daily integral of the RMM for the six countries we analyzed, against the daily RCO. These plots appear in Figure 6 and the hypothesized linear relationship is clear.

#### Additional properties of the CSIR solution

Substituting the values for *R*(*t*) and *N*(*t*) from equations S1-68 and S1-65 into equation S1-73, we can arrive at an expression for *ρ*(*t*) as a function of time:

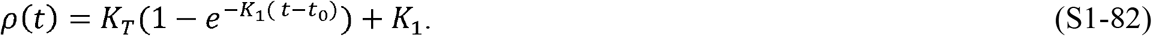

From equation S1-27, we can see that the number of infections, *I*(*t*), will begin to decrease when 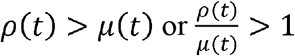. Using the previously developed expressions for *ρ*(*t*) and *μ* (*t*), we can write the following criteria for when the epidemic will begin to decline:

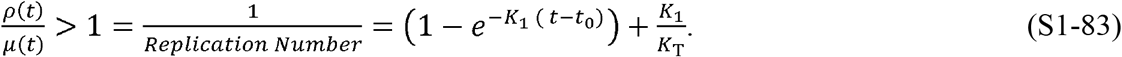

Using equation S1-83, we obtain the following expression for when the decline begins:

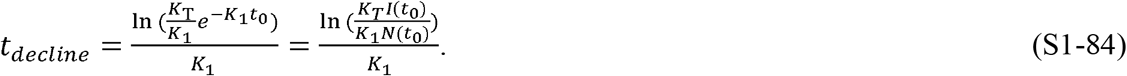

If we differentiate both sides of equation S1-64, we obtain an expression identical to equation S1-84:

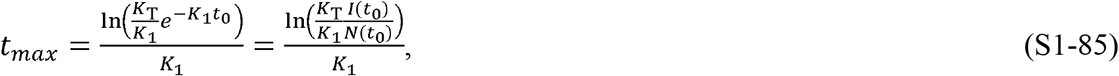

where *t*_*max*_ = the time to the peak of new infections. The time of the peak in new cases coincides, as it should, with the start of the decline of infections.

Equation S1-85 demonstrates the relationship between the strength of social intervention measures, *K*_1_, and the time to the peak of new infections. When social interventions are stronger (larger *K*_1_), the time to the peak will be shorter. Note that all increases in *K*_1_ decrease the time to the peak. The curve simply does not “flatten”; it peaks earlier and then falls steeply.

Another important expression is the rate of acceleration of the epidemic:

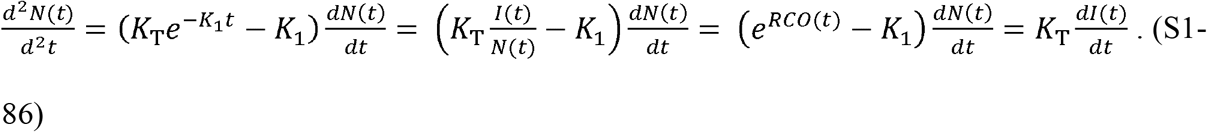

Equation S1-86, with its four equivalent expressions, is a demonstration of the power that an authentic model provides. The leftmost expression allows us to compare the acceleration—the potential to change the rate of new infections—at any stage of the epidemic for any two countries, even those with different population densities, using only the critical coefficients and the daily case rate. Equation S1-86 is an immediate determinant of whether the control measures in place, represented by *K*_1_, are effective enough. If the value of the term 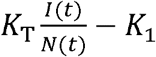 is positive, then the control measures are not strong enough. Conversely, when this term is negative, the epidemic is being brought under control.

The furthest right-hand equality in equation S1-86, 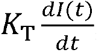, underscores that the goal of all containment measures should be to limit the creation of new infections as fast as possible. When there are more daily new infections than daily recoveries, the epidemic is accelerating and expanding. Conversely, when there are fewer daily new infections than daily recoveries, the epidemic is slowing and will end, provided the containment measures are kept in place long enough to extinguish the outbreak.

The minimum value of *K*_1_ that will begin to bring down the new cases per day occurs when the acceleration is less than zero. If we set the left-hand side of equation S1-86 to zero, and use the third expression from the left and solve for *K*_1_, we arrive at the defining relationship for this critical parameter of pandemic management:

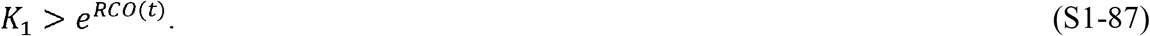

Since the value of *RCO* (*t*) is easily determined every day during the pandemic, the minimum value of *K*_1_ needed to reduce the number of daily cases can always be determined. This, in turn, determines the maximum level of infectable social contact allowable to continue to decrease the number of new daily cases.

Yet another important relationship can be derived from equation S1-64. In that equation, the term 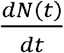 is the rate of new cases and, in Figures 1B, D, F, H, and Figure 4, this is the new cases per day. If we define a desired target for the number of new cases per day at a future time, *t* + *t* _*target*_, then we can define a new quantity, the desired fraction of the current new cases, *D*_*tf*_, as:

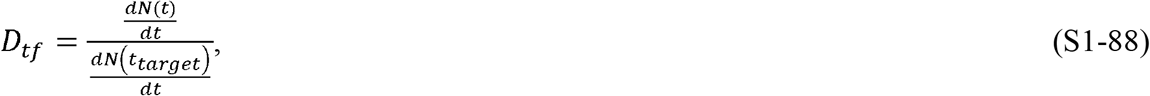

and using equations S1-64 and S1-65, we arrive at the following expression:

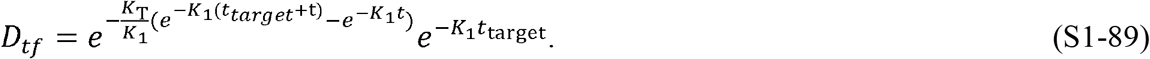

If *t* ≫ *t*_*target*_, then 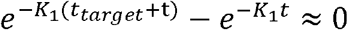 and we can derive equation S1-90 from the remaining terms:

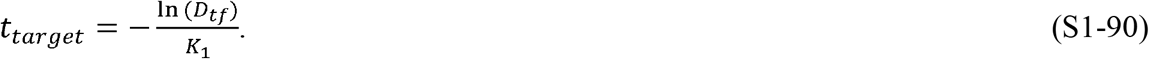

Equation S1-90 quantitates the number of days, *t*_*target*_, that a level of social containment, *K*_1_, will be required to achieve a fraction of daily cases, *D*_*tf*_, compared to the current level.

### SUPPLEMENT 2

#### Controlling epidemics early

The quantitative mathematical relationships derived from the CSIR solution in Supplement 1 characterize the dynamics of an epidemic and illustrate that strong and early intervention is critical. Equation S1-66 quantitates that the ultimate number of individuals infected in an epidemic, *N*_∞_, will be exponentially dependent on the number of people with which each person interacts.

The country data provide vivid examples. Both South Korea and New Zealand enacted strong and early interventions compared to other countries (3, 4). This is reflected in their *K*_*1*_ values (Table 1). These strong interventions led to earlier peaks in new cases and to far fewer cases than in other countries (Figure 1C–F): the peak number of new cases in both South Korea and New Zealand was 90–99% lower than in other countries, a compelling validation of the explicit statement in the CSIR solution that strong intervention leads to *exponentially* more favorable outcomes.

In the USA, intervention began to have an effect around March 23 (Figure 4B); the number of active cases on that date was 46,136 (Table 1). Using the values of *K*_*1*_ and *K*_*2*_ in Table 1, equation S1-66 predicts that the ultimate number of cases should have been approximately 1.22 million. If the same intervention had been implemented and sustained starting on March 10, when there were 59 times fewer (782) cases (7), the model predicts that the ultimate number of cases would also have been 59 times lower, or 20,725. Earlier action could have reduced the ultimate number of projected cases by more than 98%. Of course, the projected estimate of ∼1.22 million total US cases would only have occurred if the effectiveness of the interventions that were launched on March 16 had been sustained. Unfortunately, a marked reduction in effective interventions occurred widely in the USA in mid-April, well before the official reopening of the economy (13). This caused a second surge in new cases in late April and is the reason for the divergence between the observed data and the model prediction in Figure 3B. A third surge followed additional reopening activities in early fall.

As shown in Supplement 1, the CSIR solution provides an estimate of the time to the peak of new cases, *t*_max_. Using equation S1-85 and the values of *K*_*1*_ and *K*_*2*_ from Table 1, the predicted peak in new cases in the USA would have occurred near March 24 if the intervention had begun on March 10. Instead, delaying effective intervention in the USA for 16 days shifted the initial peak to April 11, 16 days later, as projected; and that peak was much higher (Figure 4B).

As shown, too, in Supplement 1, epidemic acceleration, the instantaneous potential to change the pace of the epidemic, can be determined at any point in the epidemic and depends on the social containment actions in effect at that time (equation S1-86). What is, perhaps, less apparent, but predicted by the model, is that two countries with identical numbers of cases on a given day can, in fact, have different accelerations on the same day, and exhibit different dynamics immediately after that day.

South Korea and New Zealand (Figure 2A and F) had nearly identical case counts when each imposed strong containment measures (204 cases in South Korea on February 21, and 205 in New Zealand on March 25). Their interventions model as being about equally effective (*K*_*1*_ = 0.24 in South Korea and 0.17 in New Zealand; Table 1). However, since South Korea has a much higher population density than New Zealand (Table 2), it had a much higher number of interactions when the interventions were imposed and, therefore, a higher rate of acceleration as evidenced by its higher RCO at the time of intervention. Indeed, the rate of change of new cases *was* higher in South Korea than in New Zealand, and the later number of cases in South Korea was higher than in New Zealand (Figure 4A and F).

Equation S1-83 clearly illustrates these lessons. As social distancing is strengthened (lower and therefore higher *K*_*1*_), the replication number decreases and the epidemic slows. Early and strong interventions, especially in countries with indigenously high levels of social interaction, are necessary to stop an epidemic in the initial stages; and reopening actions, enacted too early, can reignite the epidemic, dramatically increasing the number of cases. The astonishing magnitude of the effects, driven by only a few days of delay, derive from the doubly exponential nature of the underlying relationships.

### SUPPLEMENT 3

#### Ending an ongoing epidemic

The CSIR solution can also be used to design measures to end an epidemic in an advanced stage. The management plan is built by first using equation S1-90 to predict how many days a given level of intervention, *K*_1_, takes to reduce the new daily cases by a target fraction:

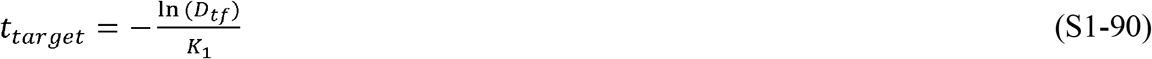

Where *t*_*target*_ is the time to the desired reduction and *D*_*tf*_ is the target as a fraction of the current level of new cases per day.

For example, if a country were to target a 90% reduction of new cases per day (e.g., from 50,000 to 5,000 cases per day, *D* _*tf*_ = 0.1), this level can be attained in about 12 days by imposing a containment level of *K*_1_= 0.2. The New Zealand and South Korea data demonstrate that equation S1-90 is valid and that *K*_1_= 0.2 is achievable for this duration. Both achieved a value of *K*_1_ close to 0.2 for the time necessary to produce a 90% reduction. It was 13 days in South Korea and 15 days in New Zealand (March 3–16, South Korea; April 2–15, New Zealand (7)).

Since 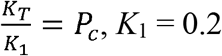, characterizes a lockdown in which people in the country can each only have one plausibly infectious contact with a little over one specific person for the containment duration. This does not mean they cannot contact anyone other than the one person; but they must use care, using masks and proper distancing, to ensure there is no plausibly infectious contact with anyone other than the one person.

Returning to the planning example: once the initial 90% reduction is achieved, a reasonable next step might be to relax the social containment to a level that allows the economy to remain viable while preventing the epidemic from erupting again. The level of necessary *K*_1_ to achieve a chosen target can be again found using equation S1-90. If an additional 90% reduction in new cases per day is desired, and a period of 90 days is tolerable for that reduction, then the new level of *K*_1_ needed is ∼0.025. This equates to allowing each person to be in contact with 7 specific people, in an infectable way, for 90 days. Note that this is 3 times *less* stringent than the original US shutdown level in April 2020. Thus, with a well-planned approach, a country can reduce its new daily cases by 99% in approximately 100 days, enabling the country to control, and essentially end, the epidemic while maintaining economic viability.

If even *K*_1_ = 0.025 is too restrictive, a lower *K*_1_ can be chosen, but it must be large enough to avoid a new outbreak. A bound for the new value of *K*_1_, low enough to prevent an outbreak and yet continue decreasing the new cases per day, can be found using equation S1-87.

The progress of interventions is easily monitored using the RCO, as the curve for South Korea illustrates (Figure 2A). Had this country maintained the implemented level of distancing measures, the data would have followed the initial slope. However, the actual data departed from the slope, heralding the failures in (or relaxations of) social distancing, which were later documented to have occurred during the indicated time frame (3) (circled data, Figure 2A).

Because it summarizes the epidemic dynamics, the RCO can be used to continuously determine if implemented measures are effective or need to be altered.

#### Outbreaks

Thus far in the development of the CSIR solution, *K*_1_ has been assumed to be constant over time. The solution is readily extended to allow *K*_1_ to vary with time in a piecewise manner. Here, we develop equations describing the analogous model in which *K*_1_ is constant in finite intervals of time, between which it changes. This addresses scenarios in which the population interactions change at certain points in time, but remain constant, or nearly so, in the intervals between those changes. In addition, because this is a common happenstance, we develop an expression for *K*_2_ for use in equation S1-65 when the model is fit to data that does not start at time *t* = 0.

If *K*_1_ and *K*_2_ are functions of time, equation S1-64 can be rewritten as

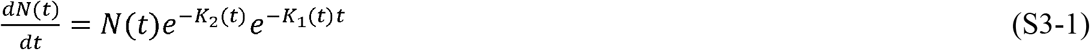

and equation S1-65 as

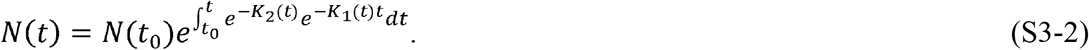

Distancing measures tend to be constant for many days at a time, so for this analysis, we assume *K*_1_ is piecewise linear. Therefore, we can calculate *K*_2_ (*t*) for any time, *t*_*n*_, when *K*_1_ changes:

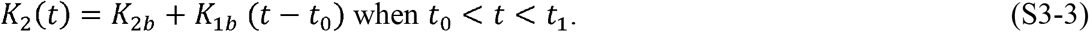

If *t*_0_ = 0, then *K*_1*b*_ and *K*_2*b*_ are the baseline levels of these parameters, and 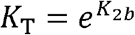. These three *K* values represent the epidemic dynamics during the initial stages, before any containment measures are implemented.

As time passes, *K*_2_ becomes

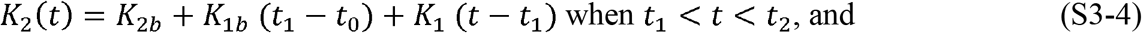

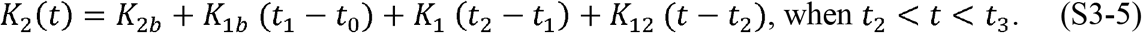

Noting the pattern as *t* increases, we can rewrite equations S3-4 and S3-5 as

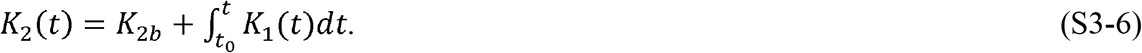

For any interval, *t*_*n*_ < *t* < *t*_*n*+1_ where *K*_1_ is a constant, *K*_2_(*t*) can be expressed as

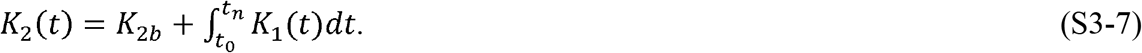

Equations S3-1 and S3-2 can also be rewritten as

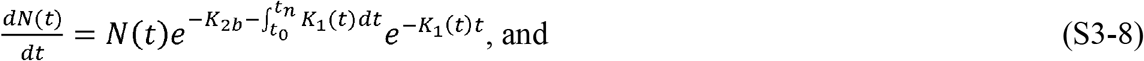

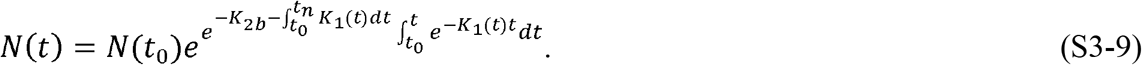

An expression for what happens in an epidemic *before* and *after* social distancing measures are implemented can be especially useful. A typical, perhaps worst-case, scenario might be “reopening”, in which social distancing measures are withdrawn and social intercourse returns to normal levels. In that instance, *K*_1_(*t*) becomes *K*_1*b*_ at time *t*_n_ when distancing measures are lifted. Using equation S1-64, we arrive at the following expressions:

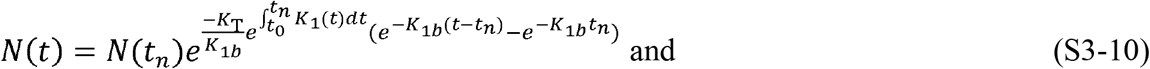

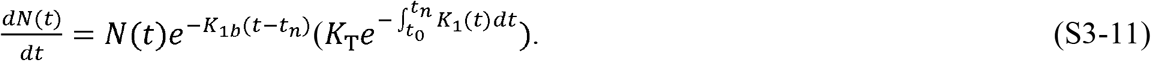

Equations S3-10 and S3-11 predict the progression of the epidemic before and after interventions are instituted. They demonstrate that the dynamics of the epidemic will depend on prior containment measures, as shown by the appearance of the expression 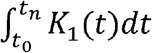 in both equations. This means that as long as the initial three assumptions of: 1) immunity; 2) no new infections are introduced from outside the area; and 3) the epidemic remains contiguous are still valid, then when interventions are relaxed, the epidemic can still grow nearly exponentially, but the growth of such an outbreak will not be as rapid as the initial outbreak. This is a consequence of the fact that, under the three assumptions, *K*_1_> 0. Since *K*_1_ is proportional to the inverse of the effective area traversed by an individual, it is proportional to the number of social interactions, and social interactions can never be less than zero. We label an outbreak when the three assumptions remain valid, but decreases due to more social interaction, a Type 1 outbreak. A Type 1 outbreak appears to have occurred in mid-April in the USA.

If new infections are introduced into a portion of the population that has been thus far disconnected from the previously infected area, then the assumption of contiguity has been violated. This is a common situation when infected people travel from an infected area into an area that was previously uninfected or had not yet seen significant numbers of infections. We label this a Type 2 outbreak.

Equation S1-65 must be modified to predict the number of cases in an epidemic that is having Type 2 outbreaks. Assuming that *t*_0_ = 0, *N* (*t*_0_) = 1, and introducing the notation *K*_1*x*_, where *x* denotes the number of the outbreak, equation S1-65 can be written as:

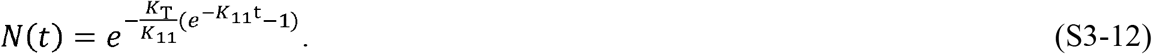

If a new outbreak occurs in a previously unaffected area of a country, then equation S1-65 can be modified as follows:

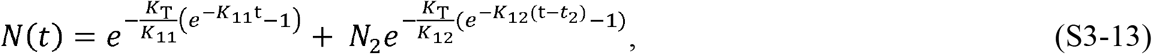

where *N*_2_ is the number of infectious people who initiated the new outbreak, *K*_12_ is the social interaction parameter in the new outbreak area, and *t*_2_ is the time the new outbreak occurs.

Equation S3-13 can be written in a general form as:

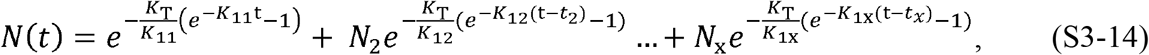

where *x* denotes the outbreak number and *t*> *t*_2_ > *t*_3_ > … > *t*_*x*_. For each outbreak, *t*_*x*_, *K*_1x_, and *N*_*x*_ need to be determined independently.

While an epidemic is underway, a Type 2 outbreak can be detected by monitoring the slope of the RCO curve. If a positive slope is detected in an RCO curve, a Type 2 outbreak has occurred. This is an indication that immediate action, within days, is required from policy makers to strengthen intervention measures and prevent the outbreak from overwhelming prior progress in controlling the epidemic.

If the disease changes its transmissibility through mutation, this can also be detected by monitoring the RCO curve. In this situation, a proper fit of the parameters in equation S1-71 will not be possible and a modification of *K*_*T*_ will be required to accommodate the change.

## FOOTNOTES

### Abbreviations

ASIR: Approximate SIR
CSIR: Complete SIR
RCO: Rate of change operator
RMM: Residential mobility measure
SIR: Susceptible–Infectious–Recovered

